# Multicentre evaluation of two multiplex PCR platforms for the rapid microbiological investigation of nosocomial pneumonia in UK ICUs: the INHALE WP1 study

**DOI:** 10.1101/2020.11.04.20216648

**Authors:** Virve I Enne, Alp Aydin, Rossella Baldan, Dewi R Owen, Hollian Richardson, Federico Ricciardi, Charlotte Russell, Brenda O. Nomamiukor-Ikeji, Ann Marie Swart, Juliet High, Antony Colles, Julie A Barber, Vanya Gant, David M Livermore, Justin O’Grady, and the INHALE WP1 Study Group

## Abstract

**Background:** ICU patients with hospital-acquired or ventilator-associated pneumonia (HAP or VAP) have high mortality, so broad-spectrum antibiotics are initiated at clinical diagnosis, then refined after 2-3 days, once microbiology results become available. Unfortunately, culture-based microbiological investigation is also insensitive, with aetiological agents remaining unidentified in many cases. This leads to extended over-treatment of patients with susceptible pathogens, whilst those with highly-resistant pathogens are treated inadequately for prolonged periods. Using PCR to seek pathogens and their resistance genes directly from clinical samples may improve therapy and stewardship. The INHALE study compared two PCR platforms for HAP/VAP diagnosis against routine microbiology (RM), identifying one to progress into a Randomised Controlled Trial (RCT).

**Methods:** Surplus routine sputa, endotracheal tube exudates and bronchoalveolar lavages were collected from ICU patients about to receive new or changed antibiotics for hospital-onset lower respiratory tract infections at 15 UK hospitals. Samples were tested (or frozen for testing) within 72h of collection. Testing was performed using the BioFire FilmArray Pneumonia Panel (bioMérieux) and Unyvero Pneumonia Panel (Curetis). Agreement between machine- and RM-results was categorised as ‘full positive/negative concordance’, ‘partial concordance’ or ‘major/minor discordance’. Bayesian latent class (BLC) analysis was used to estimate the sensitivity and specificity of each test, incorporating information from both PCR panels, 16S rDNA analysis and RM.

**Findings:** In 652 eligible samples; PCR identified pathogens in considerably more samples compared with RM: 60.4% and 74.2% for Unyvero and FilmArray respectively *vs*. 44.2%. Both tests also recorded more organisms per sample than routine culture, with the two PCR tests frequently in agreement with each other. For common HAP/VAP pathogens, FilmArray had sensitivity of 91.7-100.0% and specificity of 87.5-99.5%; Unyvero had sensitivity of 83.3-100.0%% except for *Klebsiella aerogenes* (50.0%) and *Serratia marcescens* (77.8%), and specificity of 89.4-99.0%. BLC analysis indicated that, compared with PCR, RM had low sensitivity, ranging from 27.0% to 69.4% for common respiratory pathogens. PCR detected more high-consequence antimicrobial resistance genes than would have been predicted by RM and susceptibility testing; around half the host strains could be detected when culture was repeated and they were sought assiduously.

**Interpretation:** Conventional and BLC analysis demonstrated that both platforms performed similarly and were considerably more sensitive than RM, detecting potential pathogens in patient samples reported as culture negative. FilmArray had slightly higher sensitivity than Unyvero for common pathogens and was chosen for INHALE’s RCT, based on the balance of these results, a swifter turnaround time (75 min vs. 6h), and a smaller footprint. The increased sensitivity of detection realised by PCR offers potential for improved antimicrobial prescribing.

## Introduction

Pneumonia is the most-frequently-reported infection in intensive care unit (ICU) patients, responsible for significant morbidity and mortality worldwide.^1-3^ It is differentiated into community-acquired pneumonia (CAP, originating outside of the hospital or within 48h of admission), hospital-acquired pneumonia (HAP, developing >48h after hospital admission) and ventilator-associated pneumonia (VAP, developing in patients mechanically ventilated for at least 48h).^4^ Estimates of crude mortality for nosocomial pneumonia (HAP and VAP) range from 30-70%.^1^ Administration of active antimicrobial therapy as soon as possible after clinical onset is crucial for a successful outcome, whereas inadequate treatment – meaning delayed antibiotics or those that transpire not to be active – increase mortality.^5,6^

The various bacteria, viruses and (rarely) fungi that cause nosocomial pneumonia cannot be distinguished from clinical symptomology; instead microbiological diagnosis is needed, along with antibiotic susceptibility testing for bacteria. Results do not become available for at least 48-72h after a respiratory sample is taken, forcing clinicians to treat empirically in the interim. The diversity of nosocomial bacteria that can cause HAP and VAP means that broad-spectrum antibiotics must be used, as indicated in current EU, US and UK guidelines. ^1,3 7,8^

The current gold-standard for aetiological investigation is microbiological culture, hereafter termed routine microbiology (RM). Evidently, this can only grow cultivable bacteria and depends on these being recoverable from the sample. In up to 75% of pneumonia cases culture fails to identify a bacterial pathogen, even though many of these cases are believed to be bacterial and continue to be treated with antibiotics on this basis.^9-11^ This combination of slow turnaround and poor sensitivity results in the continued administration of broad-spectrum therapy rather than switching to pathogen-targeted narrow-spectrum antibiotics. This amounts to poor stewardship and may increase the risk of side effects, including selection of *Clostridium difficile* disease, and driving the emergence of antibiotic resistance in the gut flora.^12^ A further hazard, particularly in high-resistance countries, is that even the empirical broad-spectrum agent proves ineffective against the pathogen, increasing the risk of a poor clinical outcome.

Rapid and accurate diagnostics provide a potential route to improving this unsatisfactory situation, providing scope for early refinement of individual patients’ therapy and potentially improving clinical outcomes as well as antimicrobial stewardship. Commercial “sample-in, answer-out” PCR-based pneumonia tests have become available, including the Unyvero (Curetis) and BioFire FilmArray (bioMérieux) platforms. These are substantially automated and seek prevalent pathogens and critical resistances. They require minimal hands-on time and allow turnaround within hours instead of days, potentially reshaping clinical microbiology and the management of pneumonia patients.^13-16^

The aim of this study was to evaluate and compare the performance, in terms of pathogen and resistance marker detection, of the Unyvero Hospitalized Pneumonia (HPN) Cartridge and BioFire FilmArray Pneumonia Panel, using lower respiratory tract samples from patients clinically diagnosed with HAP or VAP at 15 ICUs in the UK. Machine results were compared with each other, RM and 16S rRNA sequencing. The analysis also sought to choose one test to take forward into a randomized controlled trial (RCT), now underway, to evaluate its clinical utility.

## Materials and Methods

### Patients and specimens

Between September 2016 and May 2018, surplus routine lower respiratory tract samples were collected from eligible patients with suspected HAP/VAP across the 15 ICUs participating in the INHALE study. These sites were selected in order to represent a range of UK institution types, and included tertiary referral centres, district general hospitals, one children’s hospital and one private hospital. They comprised Aintree University Hospital NHS Foundation Trust, Bupa Cromwell Hospital, Chelsea and Westminster Hospital NHS Foundation Trust, City Hospitals Sunderland, Dudley Group NHS Foundation Trust, Great Ormond Street Hospital, Guy’s and St Thomas’ NHS Foundation Trust, James Paget University Hospitals NHS Foundation Trust, Norfolk and Norwich University Hospitals NHS Foundation Trust, North Middlesex University Hospital NHS Trust, Queen Elizabeth Hospital Kings Lynn NHS Trust, Royal Free Hospital, Royal Liverpool and Broadgreen University Hospitals NHS Trust, University College London Hospitals and University Hospitals of North Midlands and they were served by 11 different microbiology laboratories

Specimens were included if they were of sufficient volume (>400 μl) and were from ICU inpatients hospitalized for at least 48h who were about to receive a new antibiotic or change in antimicrobial therapy for suspected lower respiratory tract infection. Specimens were eligible for inclusion only when collected from the patient within 12h (before or after) of antimicrobial therapy being administered and then tested, or frozen at −80°C, within 72h of collection. All types of lower respiratory specimens were accepted, but upper respiratory tract specimens (e.g. nasopharyngeal aspirates) were excluded. Second specimens from the same patient were included only when collected > 14 days after the first sample.

### Ethical approval

This work had study specific approval from the UK Health Research Authority (Reference: 16/HRA/3882, IRAS ID: 201977,) and the UCL DNA Infection Bank Committee whose operation is governed by the London Fulham Research Ethics committee (REC Reference: 17/LO/1530).

### Conventional culture and susceptibility testing

Each respiratory specimen was initially cultured locally, at the laboratory serving the participating hospital, according to their standard operating procedures (SOPs). These SOPs were all based on the Public Health England (PHE) UK Standard.^17^ Except in the case of bronchoalveolar lavage (BAL) specimens this specifies initial homogenisation of the respiratory sample with 0.1% dithiothreitol, followed by a 10^−5^ dilution, and inoculation of the diluted and undiluted specimen onto chocolate agar with bacitracin (incorporated or as a disc), cysteine lactose electrolyte deficient agar (CLED) or MacConkey agar, and Sabouraud agar for fungi. Blood agar may also be added, especially if bacitracin is incorporated in the chocolate agar. In the case of Bronchoalveolar lavage (BAL), culture is performed on serial dilutions of a sample that has been concentrated by centrifugation.

Plates are incubated at 35-37°C in the presence of 5% CO_2_ (blood and chocolate agar) or in air (MacConkey and CLED agar) for 40-48h, with daily reading of results. Bacterial pathogens are identified to species level by MALDI-TOF or biochemical methods, followed by antimicrobial susceptibility testing using EUCAST or BSAC interpretive standards.

The PHE standards provide guidance on the interpretation of culture results for BAL samples, but interpretation and reporting are left to the discretion of individual laboratories for other sample types.

### PCR Testing

Samples were transported to the two central research laboratories (University of East Anglia and University College London, Royal Free Hospital campus) by courier. Upon receipt, each specimen was promptly tested using both the Unyvero Pneumonia Panel (Curetis, Holzgerlingen, Germany) and the BioFire FilmArray Pneumonia Panel (BioFire Diagnostics, Salt Lake City, USA) according to manufacturer’s instructions. The basic characteristics of each test, along with the panel of targets sought are detailed in Table 1.

**Table 1.** Features and Target Panels of the Curetis Unyvero Pneumonia Panel and the BioFire FilmArray Pneumonia Panel multiplex PCR tests.

|  |  |  |
| --- | --- | --- |
| Characteristic | Curetis Unyvero HPN Hospitalised Pneumonia Panel | BioFire FilmArray Pneumonia Panel *plus* |
| Technology | Automated sample preparation, multiplex PCR and microarray detection of targets | Automated sample preparation and nested PCR |
| Regulatory status | CE-IVD<sup>1</sup> | CE-IVD & FDA Cleared<sup>2</sup> |
| Hands-on preparation time | 2 min | 2 min |
| Run-time | 5h | 1h 15 min |
| Bacteria sought | *Acinetobacter baumannii* complex<br>*Citrobacter freundii*<br>*Enterobacter cloacae* complex<br>*Escherichia coli*<br>*Haemophilus influenzae*<br>*Klebsiella aerogenes*<br>*Klebsiella oxytoca*<br>*Klebsiella pneumoniae*<br>*Klebsiella variicola*<br>*Moraxella catarrhalis*<br>*Morganella morganii*<br>*Proteus* spp.<br>*Pseudomonas aeruginosa*<br>*Serratia marcescens*<br>*Stenotrophomonas maltophilia*<br>*Streptococcus pneumoniae* | *Acinetobacter calcoaceticus-baumannii* complex<br>*Enterobacter cloacae* complex<br>*Escherichia coli*<br>*Haemophilus influenzae*<br>*Klebsiella aerogenes*<br>*Klebsiella oxytoca*<br>*Klebsiella pneumoniae*<br>*Moraxella catarrhalis*<br>*Proteus* spp.<br>*Pseudomonas aeruginosa*<br>*Serratia marcescens*<br>*Streptococcus agalactiae*<br>*Streptococcus pneumoniae*<br>*Streptococcus pyogenes* |
| Atypical organisms | *Chlamydophila pneumoniae*<br>*Legionella pneumophila*<br>*Mycoplasma pneumoniae* | *Chlamydophila pneumoniae*<br>*Legionella pneumophila*<br>*Mycoplasma pneumoniae* |
| and Fungi<br>sought | *Pneumocystis jirovecii* |  |
| Viruses<br>sought | None | Adenovirus<br>Coronaviruses OD43, NL63,<br>HKU1 and 229E<br>Human metapneumovirus<br>Human rhinovirus/enterovirus<br>Influenza A<br>Influenza B<br>Parainfluenza virus<br>Respiratory syncytial virus<br>MERS Coronavirus |
| Antimicrobial<br>Resistance<br>Genes<br>sought | *ermB*<br>*mecA*<br>*mecC*<br>*bla*<sub>TEM</sub><br>*bla*<sub>SHV</sub><br>*bla*<sub>IMP</sub><br>*bla*<sub>KPC</sub><br>*bla*<sub>NDM</sub><br>*bla*<sub>OXA-23</sub><br>*bla*<sub>OXA-24/40</sub><br>*bla*<sub>OXA-48</sub><br>*bla*<sub>OXA-58</sub><br>*bla*<sub>VIM</sub><br>*sul1*<br>*gyrA83*<br>*gyrA87*<br>*mecA/C* and MREJ | *bla*<sub>KPC</sub><br>*bla*<sub>NDM</sub><br>*bla*<sub>OXA-48</sub> like<br>*bla*<sub>VIM</sub><br>*bla*<sub>IMP</sub><br>*bla*<sub>CTX-M</sub> |

<sup>1</sup>A similar panel, featuring a reduced number of antimicrobial resistance genes, has FDA clearance.
<sup>2</sup>We evaluated the Research Use Only (RUO) version

### Supplementary analyses

All specimens with a sufficient surplus (300 µl) after PCR testing underwent 16S metagenomic analysis. Samples were inactivated by incubating for 30 minutes at 99°C, then DNA was extracted using the ZR Viral RNA/DNA kit and ZR BashingBead Lysis Tubes (Zymo Research). Briefly, 300 µl of sample were transferred in a bead tube, homogenized in a bead-beater for 30 seconds at 3,500 oscillations per minute, centrifuged for 1 minute at 21,000 g. Next, 200 µl of supernatant were transferred to a clean Eppendorf tube and DNA was extracted following manufacturer’s instructions. Illumina 16S metagenomic sequencing was then performed according to the manufacturer’s protocol (Illumina, 15044223B): the V3-V4 16S rRNA region was amplified on a LightCycler 480 II instrument (Roche) and sequenced on an Illumina MiSeq system. The Illumina BaseSpace 16S metagenomic pipeline was used to analyse the results. Only samples with at least 10,000 total reads were deemed eligible for analysis. For a genus to be considered significant, it had to comprise at least 1% of all reads.

A sub-set of specimens, selected at random among those where culture and PCR disagreed for resistance detection underwent additional culture-based analysis, termed comprehensive culture (CC) at the UCL research laboratory, using methodology described previously.^15^ Briefly, a sweep of growth was taken across the plate of a fresh primary culture of the specimen on chocolate agar and stored in Microbank^TM^ vials at −80°C until analysis. Ten microliters of neat sample and a 10^−5^ dilution in 0.9% saline were then plated onto chocolate agar, Columbia blood agar (CBA), Brilliance UTI agar (Oxoid, Basingstoke, UK) and Columbia colistin-nalidixic acid agar (C-CNA) (Oxoid). CBA, UTI and C-CNA plates were incubated at 37°C in air for 18h; chocolate agar plates were incubated in 5% CO_2_ at 37°C for 18h. Representative bacterial colonies of different morphologies on each medium were identified by MALDI-TOF MS (Bruker GmbH, Mannheim, Germany), either directly from colonies or by using formic acid extraction where necessary.

### Identification of antimicrobial resistances

Additional investigation of antimicrobial resistances, or the genes responsible was performed on isolates found resistant in microbiology laboratories or CC or when either of the two molecular systems detected key resistance genes.

Gram-negative bacteria (i) reported resistant to cephalosporins or carbapenems in RM, (ii) found to have ESBL or carbapenemase genes using the PCR systems, or (iii) grown in CC were tested for resistance to ceftazidime, cefotaxime, ceftriaxone, ertapenem, meropenem and imipenem (Enterobacterales) or imipenem, meropenem, ceftazidime and piperacillin/tazobactam (*Acinetobacter* spp. and *P. aeruginosa*) by EUCAST disc diffusion methodology.^18^ Potential methicillin-resistant *Staphylococcus aureus* (MRSA) were screened for resistance to cefoxitin.

When isolates had phenotypes consistent with the presence of antimicrobial resistance genes, further genetic testing was performed. Enterobacterales resistant to a carbapenem or to oxyimino cephalosporins, *P. aeruginosa* resistant to both carbapenems and cephalosporins and *A. baumannii* resistant to imipenem or meropenem were tested with the Check-MDR CTX103XL kit (Checkpoints, Wageningen, the Netherlands) according to manufacturer’s instructions, following extraction of total genomic DNA using the Qiagen DNA Mini Kit (Qiagen). *S. aureus* isolates resistant to cefoxitin underwent in-house PCR (primers and conditions described previously)^19,20^ for detection of *mecA* and *mecC* using HotStartTaq PCR Mastermix (Qiagen) on DNA extracted with the Qiagen DNA Mini Kit.

### Data collection

RM data available on the Laboratory Information Management Systems (LIMS) of each participating hospital was were collected and managed using REDCap^21^ electronic data capture tools hosted at Norwich Clinical Trials Unit. For each included sample, we collected the culture result as reported to treating clinicians and details of significant organisms reported and their full antimicrobial susceptibility profile. Any results for relevant respiratory pathogens detected by non-culture-based methods were also included. Virology data (PCR) were collected if testing had been performed on the same calendar day as collection of the lower respiratory tract sample. We also collected details required to confirm patient eligibility and times samples were collected, processed and results released. All PCR and supplementary data generated by study staff were also recorded in RedCap. All data were anonymised.

### Data Analysis

Analyses were carried out using Stata (v 15) and R (v 3.5 or above) and followed a pre-defined, detailed statistical analysis plan. Results from the conventional and PCR tests, including about timings, detection of individual pathogens, viruses and fungi were described using standard summary statistics. We considered agreement between results from RM and the PCR tests by categorising each sample in terms of overall concordance of organisms detected. Definitions of the categories are detailed in table 2. For each PCR test, the proportions in the concordance categories were calculated, with 95% confidence intervals.

**Table 2.** Concordance-based performance of PCR tests compared with RM

| Category | Definition | All Detections |  | Detections reported at higher concentrations<sup>a</sup> |  |
| --- | --- | --- | --- | --- | --- |
|  |  | Unyvero<br>(%, 95% CI) | FilmArray<br>(%, 95% CI) | Unyvero<br>(%, 95% CI) | FilmArray<br>(%, 95% CI) |
| Full positive concordance | Organisms detected were an exact match | 19.3<br>(16.2 - 22.4) | 18.2<br>(15.2 - 21.3) | 22.4<br>(19.1 - 25.8) | 21.1<br>(17.9 - 24.3) |
| Full negative concordance | No organisms detected by either method | 37.3<br>(33.4 - 41.1) | 32.1<br>(28.4 - 35.8) | 42.1<br>(38.1 - 46.0) | 44.5<br>(40.6 - 48.4) |
| Partial concordance | PCR detected the same organism as RM plus additional organism(s) | 18.2<br>(15.1 - 21.2) | 21.0<br>(17.8 - 24.2) | 11.6<br>(9.0 - 14.1) | 11.8<br>(9.2 - 14.3) |
| Minor discordance | RM was negative but machine found $\geq 1$ organism | 20.6<br>(17.4 - 23.8) | 26.9<br>(23.4 - 30.4) | 15.8<br>(12.9 - 18.7) | 14.5<br>(11.7 - 17.3) |
| Major discordance | RM found $\geq 1$ organism, at least one of which was on the PCR panel, but not detected | 4.6<br>(2.9 - 6.3) | 1.8<br>(0.7 - 2.8) | 8.1<br>(5.9 - 10.3) | 8.1<br>(5.9 - 10.2) |

CI - confidence interval
<sup>a</sup> Calculated based on semi-quantitative detections Reported as ++ or +++ by Unyvero or $10^6$ or $\geq 10^7$ copies/ml by FilmArray

For both PCR tests, sensitivity, specificity, positive predictive value (PPV) and negative predictive value (NPV) were estimated (with exact 95% confidence intervals) for each target using RM and routine virology as the gold standard. Owing to concerns that RM provides a poor gold standard (see Results), estimates (with 95% credible intervals) were also calculated using Bayesian latent class (BLC) models^22^ incorporating results from both PCR tests, 16S metagenomics and RM. Models used non-informative priors for all parameters (except to constrain specificities above 0.15 to obtain more stable posterior distributions) and were fitted with and without assuming correlation between tests. The best fitting models were identified based on Deviance Information Criteria (DIC).

### Scoring system for evaluation of PCR-based Diagnostic Tests

At the outset of the study, through expert consensus, a scoring system was developed in order to assess the suitability of each ‘sample-in, answer-out’ test for use in the INHALE randomized controlled trial and more generally, for routine diagnostic use. Tests were assessed against one essential criterion which had to be met for further evaluation, and 10 points-based ‘Desirable Criteria’, scoring a total of 150. The essential criterion stipulated that incidence of major discordances, defined as failures to detect pathogens found by RM, was less than 5%.

Desirable Criteria were: (i) overall concordance (max. 45 points), (ii) sensitivity (20 points), (iii) failure rate (15 points), (iv) breadth of panel (15 points), (v) time to result (15 points), (vi) cost per test (15 points), (vii) footprint (5 points), (viii) consumable logistics (5 points), (ix) quality of customer service (5 points) and (x) ease of use (10 points). Criteria i-iii were scored based on study results, criteria iv-viii on manufacturer’s published information and criteria ix and x on a user questionnaire. The scale was deliberately weighted towards characteristics related to detection of pathogens, with implementation-based criteria given a lower weighting. Full details of the scoring system can be found in supplementary data (Table S1).

## Results

### Specimens Collected

A total of 752 specimens, 652 of them eligible, were collected from 15 participating ICUs (Figure 1). The range of eligible samples per site was 7-141, with 9 sites providing >20 eligible samples. Most were from adults, with just 72 (11.0%) from children. Among all samples, 260 (39.9%) came from patients with suspected HAP and 392 (60.1%) from patients with suspected VAP. Endotracheal aspirates were the most common sample type, accounting for 299 samples (45.9%); followed by sputa (272 samples, 41.7%), BALs (44 samples, 6.7%) and non-directed BALs (23 samples, 3.5.%), with the remaining 14 samples in the “other” or “unknown” category. A small majority of samples were collected before antibiotic administration (357, 54.8%).

**Figure 1.**
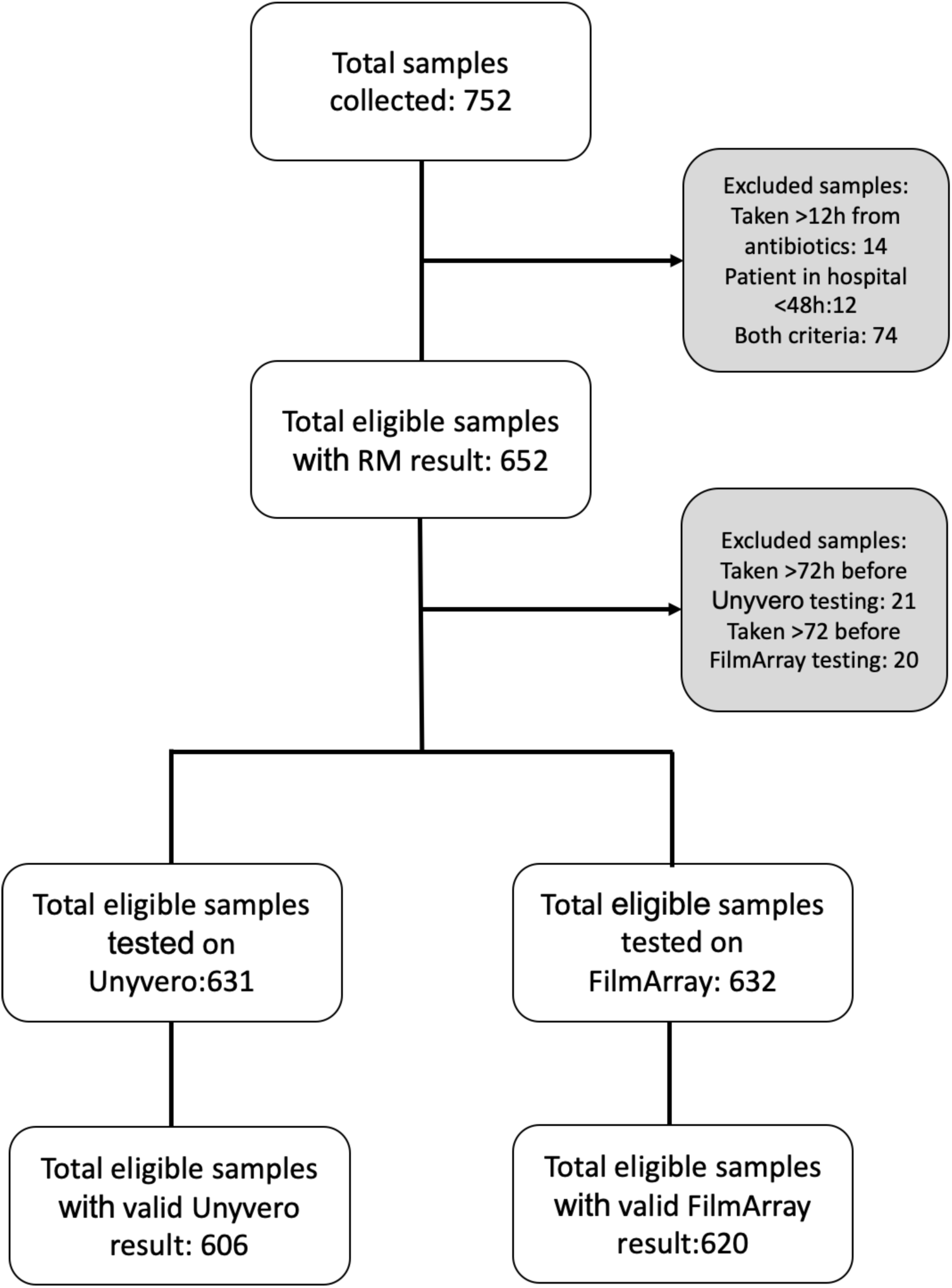
Schematic representation of sample eligibility

### Routine Microbiology Results

RM culture was performed at the local laboratories on all samples. The overall median time to a result was 70.2h (interquartile range (IQR) 51.1h to 92.1h), including a median of 6.1h (IQR 2.5h to 15.4h) transit time from the ICU to booking in at the laboratory and 55.5h (IQR 44.8h to 76.5h) from sample booking to release of results. The positivity rate was 44.2% and most positive results recorded just one significant organism (35.1%), with a minority reported as containing two or more (9.1%). The remaining 55.8% were reported mostly as ‘normal flora’, ‘non-significant growth’, or simply as no ‘growth’.

*S. aureus* was the most-frequently-found bacterium (Figure 2), representing 23.6% (83/352) of all organisms reported, closely followed by *P. aeruginosa* at 20.7 *%*; *Enterobacterales* collectively accounted for 38.1% of isolates with *Klebsiella* spp. (12.5% of all isolates) and *E. coli* (12.2%) both prominent (Figure 3a). Although *H. influenzae* is not primarily considered a hospital-acquired organism, it accounted for 6.5% of organisms cultivated. Occasionally RM laboratories reported *Candida* spp., *Enterococcus* spp. and coagulase-negative staphylococci. Because there is no evidence base for their involvement in pneumonia, they were not included in the analyses.

**Figure 2.**
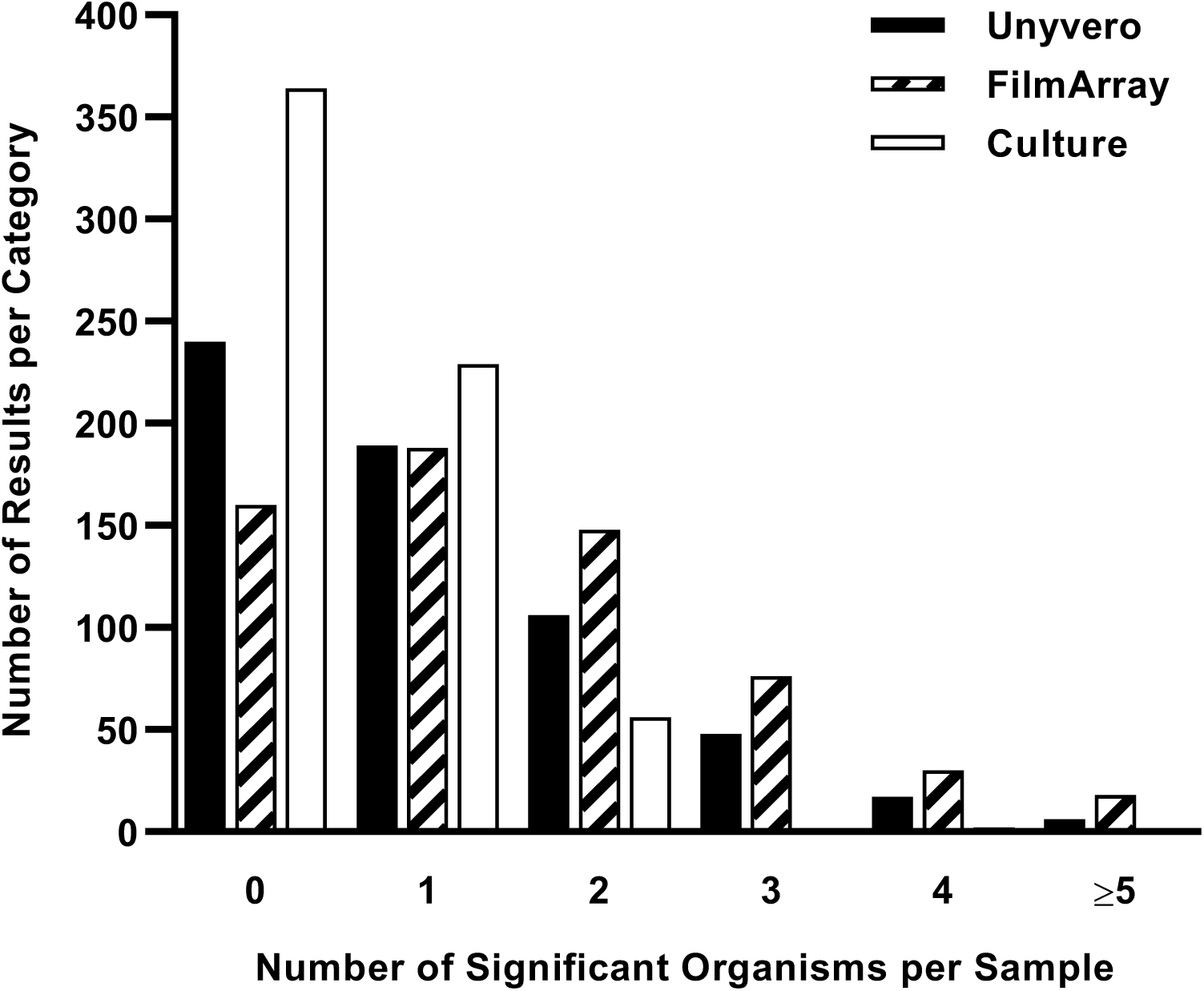
Number of significant organisms detected per respiratory sample by RM or PCR.

**Figure 3a.**
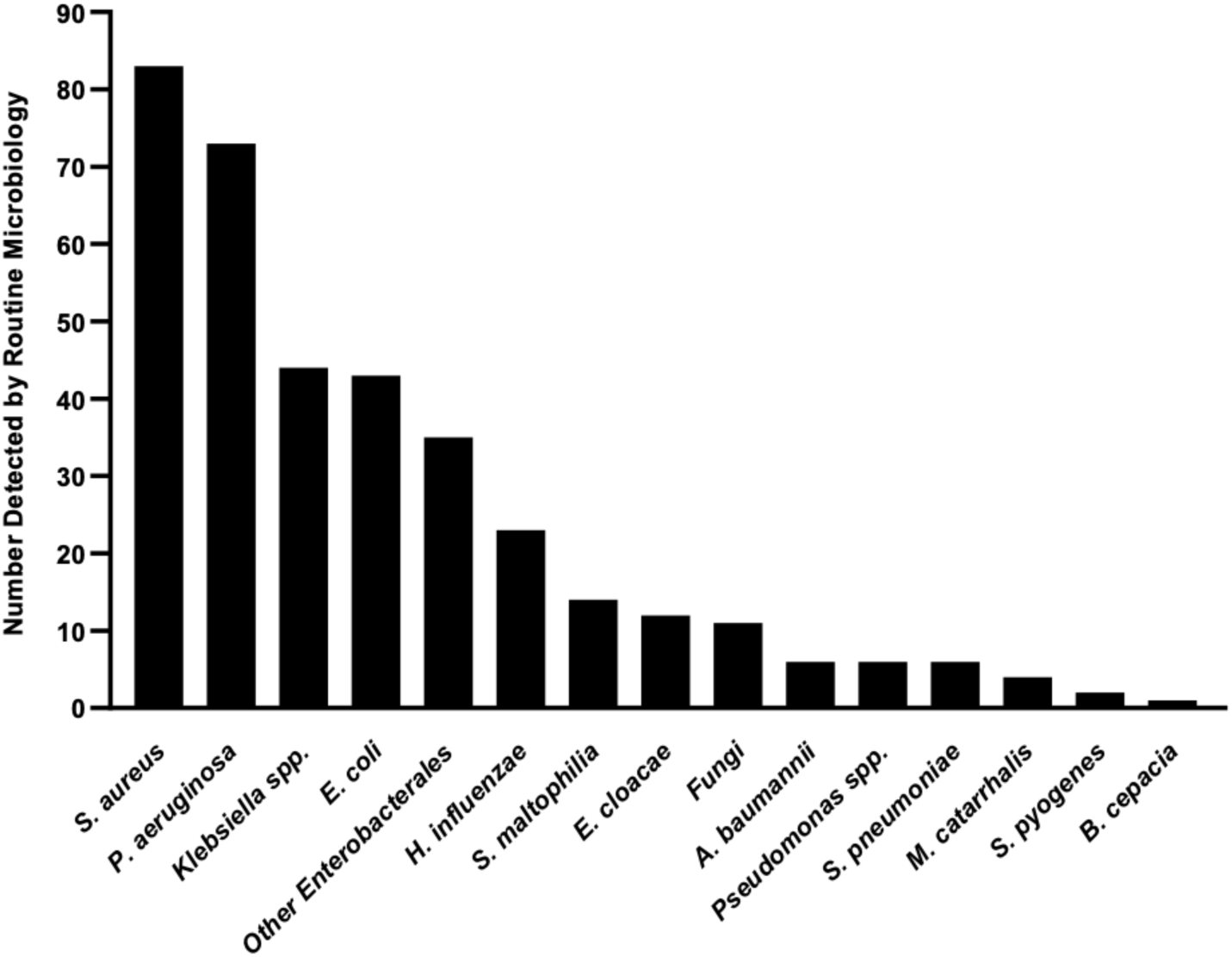
Numbers and types of bacteria detected by RM culture from respiratory samples included in the study.

Results of standard-of care for respiratory virus diagnostics were recorded if testing was performed in the same 24h period as collection of the eligible bacteriology specimen. Only 113 patients, 33 of them children, had a virology result meeting this eligibility criterion, and, of these, 31 (27.4%) were positive. The most-frequently-identified viruses were influenza A (n = 7), adenovirus (n = 6) and cytomegalovirus (n = 6).

### PCR Results

Of 652 eligible samples, 631 tested on the Unyvero system were eligible for further analysis and 632 of those tested on the FilmArray were eligible (Figure 1). The remainder were excluded owing to not being tested or frozen within 72h of collection. Among the eligible samples, 620 generated a result on the FilmArray, whilst tests on the remaining 12 failed, a test failure rate of 1.9%. Defining failure on the Unyvero is more complex since target organisms and genes are divided into 8 chambers and either the entire test or one or more chambers may fail. In the latter case, useful results may still be generated from the remaining chambers. We considered any test where >2 chambers failed as a “total failure”. In a further 32 cases just 1 or 2 chambers failed and data from the remaining 6 or 7 chambers were included in the analysis for these partial failures, with the proviso that organisms sought by the failed chambers may have been missed. A further 24 samples generated other types of total machine failure (unrelated to the chambers) which failed to generate a result for the Unyvero; altogether, there were 606 valid results, a total failure rate of 4.0%.

The overall positivity rate of both PCR tests was higher than RM, at 60.4% for the Unyvero and 74.2% for the FilmArray (chi-square test: p < 0.0001). Most specimens had multiple organisms detected (Figure 2), with this proportion higher for FilmArray than the Unyvero. FilmArray found only bacteria in 54.2% of samples and only viruses in 6.9% whereas 13.1% contained both viruses and bacteria. The most common bacterial pathogens detected by the two PCR tests are shown in Figure 3b. The principal species and their relative prevalence were broadly similar to that recorded by RM, although *E. coli* and *Klebsiella* spp. were detected more frequently in relative terms by PCR than by culture, whereas *S. aureus* and *P. aeruginosa* were detected less frequently. Between them, these four species were the most-commonly-detected organisms by both PCR and RM. Among viruses detected by the FilmArray (Unyvero doesn’t seek viruses), rhinovirus was the most prominent (n=55), followed by influenza A (n=29) and influenza B (n=25) (full viral detection data are shown is Table S2).

**Figure 3b.**
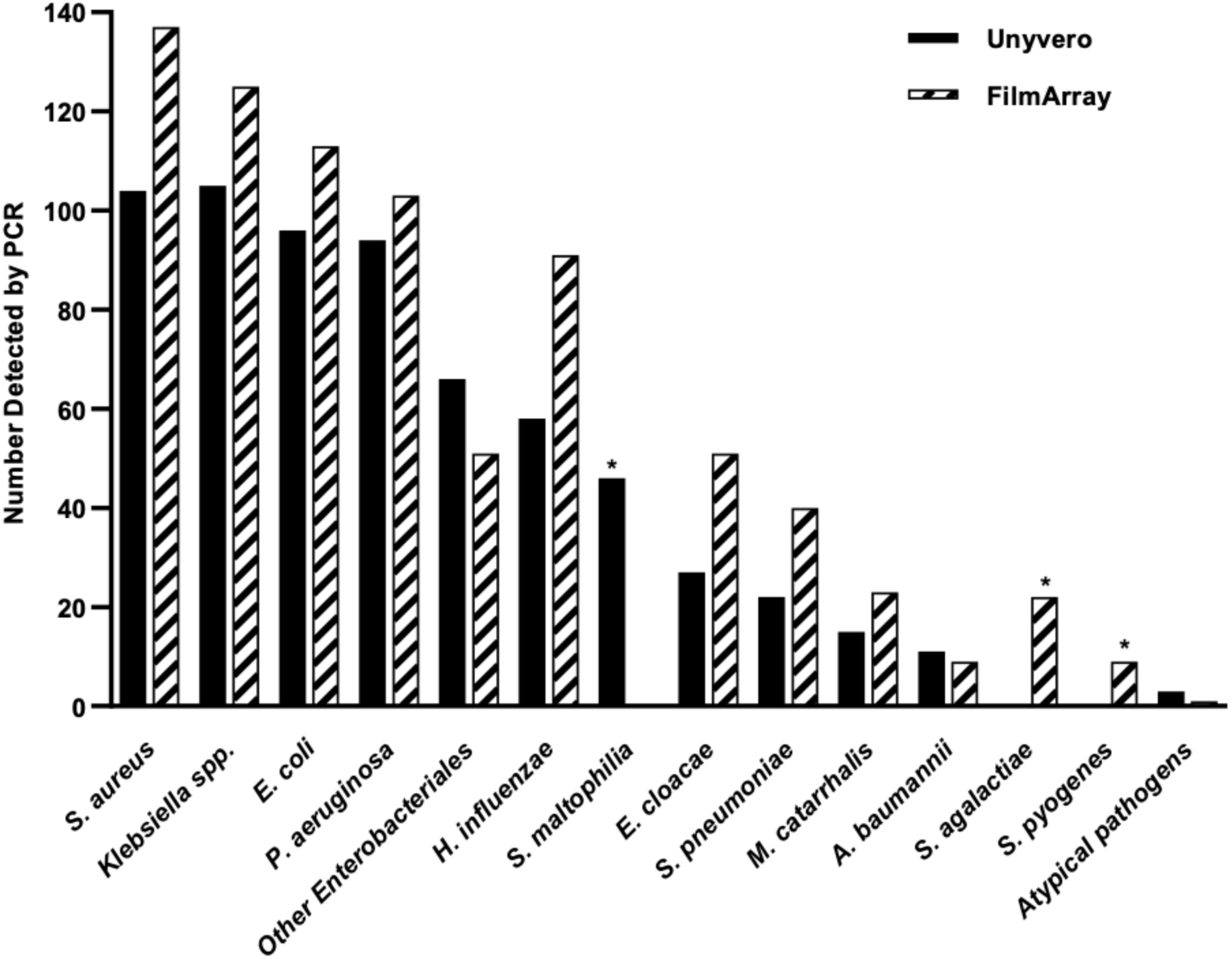
Numbers and type of bacteria detected by PCR from respiratory samples included in the study. Unyvero, solid bars, n = 606; FilmArray, hatched bars, n = 620. Species sought by one test only are marked with an asterisk.

### Performance of PCR Tests

The performance of the PCR tests was measured and compared in several ways, in order to accommodate for the fact that RM is an imperfect gold standard. Moreover, the PCR tests detect multiple targets, more than one of which may be present in any given sample. Consequently, it is challenging to calculate overall test sensitivity and specificity; it was commonplace, for example, for PCR to detect two organisms, one of which matches an organism reported by RM and the other does not.

First, overall test performance was measured in terms of concordance with RM, as shown in Table 2. Both PCR tests deliver semi-quantitative outputs; the FilmArray reports bacterial targets as 10^4^, 10^5^, 10^6^ or ≥10^7^ copies per ml, whereas the Unyvero reports pathogen targets as +, ++ or +++. We therefore further calculated concordance taking into account only those targets that were detected at high concentration, defined as 10^6^ or ≥10^7^ copies/ml for FilmArray and ++ or +++ for Unyvero. These results are also included in Table 2.

Around one half of PCR results by either method demonstrated full positive or negative concordance with RM when all PCR detections were considered. Most of the remainder were either partially concordant or had minor discordance, predicated on PCR reporting more organisms than RM. Major discordance was rare, totalling only 4.6% of Unyvero results and 1.8% of FilmArray results. If PCR detections at low concentrations were excluded, full concordance increased for both tests but major discordance also increased substantially. Target-specific sensitivity data are shown in Table 3. Local microbiology laboratories do not normally seek atypical pathogens in HAP/VAP patients, so results for *Chlamydophila pneumoniae, Legionella pneumophila* and *Mycoplasma pneumoniae* are excluded from sensitivity analyses. Unyvero and FilmArray each detected *M. pneumoniae* once, in the same specimen, where it was not sought by local microbiology. Unyvero detected two samples with *L. pneumoniae*; FilmArray and RM did not detect any. None of the three methods detected *C. pneumophila*.

**Table 3.** Pathogen-specific performance of PCR tests as compared with RM as the gold standard. 95% confidence intervals are omitted to aid readability but are included in supplementary table S3 along with frequencies of detection.

| Organism | Unyvero |  |  |  | FilmArray |  |  |  |
| --- | --- | --- | --- | --- | --- | --- | --- | --- |
|  | Sensitivity | Specificity | PPV | NPV | Sensitivity | Specificity | PPV | NPV |
| *A. baumannii* complex | 100.0 | 99.0 | 45.5 | 100.0 | 100.0 | 99.5 | 66.7 | 100.0 |
| *C. freundii* | 100.0 | 98.7 | 11.1 | 100.0 | NA** | NA | NA | NA |
| *E. cloacae* | 100.0 | 97.5 | 44.4 | 100.0 | 91.7 | 93.4 | 21.6 | 99.8 |
| *E. coli* | 87.8 | 89.4 | 37.5 | 99.0 | 97.6 | 87.5 | 36.3 | 99.8 |
| *H. influenzae* | 100.0 | 93.7 | 36.2 | 100.0 | 95.2 | 88.1 | 22.0 | 99.8 |
| *K. aerogenes* | 50.0 | 99.5 | 50.0 | 99.5 | 100.0 | 99.2 | 54.5 | 100.0 |
| *K. oxytoca* | 90.9 | 95.0 | 25.0 | 99.8 | 100.0 | 95.2 | 27.5 | 100.0 |
| *K. pneumoniae* | 83.3 | 94.2 | 37.0 | 99.3 | 92.0 | 91.4 | 31.1 | 99.6 |
| *M. catarrhalis* | 100.0 | 98.2 | 26.7 | 100.0 | 100.0 | 96.9 | 17.4 | 100.0 |
| *M. morganii* | 100.0 | 98.3 | 9.1 | 100.0 | NA | NA | NA | NA |
| *P. aeruginosa* | 95.3 | 93.9 | 64.9 | 99.4 | 98.5 | 93.1 | 63.1 | 99.8 |
| *S. aureus* | 87.2 | 93.2 | 65.4 | 98.0 | 96.2 | 88.9 | 56.2 | 98.2 |
| *S. agalactiae* | NA | NA | NA | NA | ND | 96.5 | 0.0 | 100.0 |
| *S. maltophilia* | 92.9 | 94.4 | 28.3 | 99.8 | NA | NA | NA | NA |
| *S. marcescens* | 77.8 | 98.3 | 41.2 | 99.7 | 100.0 | 98.2 | 45.0 | 100.0 |
| *S. pneumoniae* | 100.0 | 97.3 | 27.3 | 100.0 | 100.0 | 94.5 | 15.0 | 100.0 |
| *S. pyogenes* | NA | NA | NA | NA | 100.0 | 98.9 | 22.0 | 100.0 |

\*ND – not determined because RM detected no positives; \*\*NA – not applicable; organism not on test panel

Owing to the small number of routine virology results available for comparison, confidence intervals for viral detections were wide (Table S2, FilmArray only, as Unyvero does not seek viruses).

For most target bacteria, PCR assay sensitivity was >95% and NPV was > 98%. Specificity and PPV were lower, due to the PCR tests detecting more organisms per sample and finding more positive samples than RM. Notably, both machines often found the *same* organism as each other when RM did not record any organism. Such agreement is unlikely to be random and suggests RM false negative rather than PCR false positive results may represent an imperfect gold standard. Accordingly, BLC modelling was introduced, rather than gold standard comparison, to deliver a more representative comparison of all tests. Table 4 shows performance estimates obtained from BLC models that do not rely on RM as a gold standard. Based on DIC values, models that assumed independence of the test results were preferred.

**Table 4.**
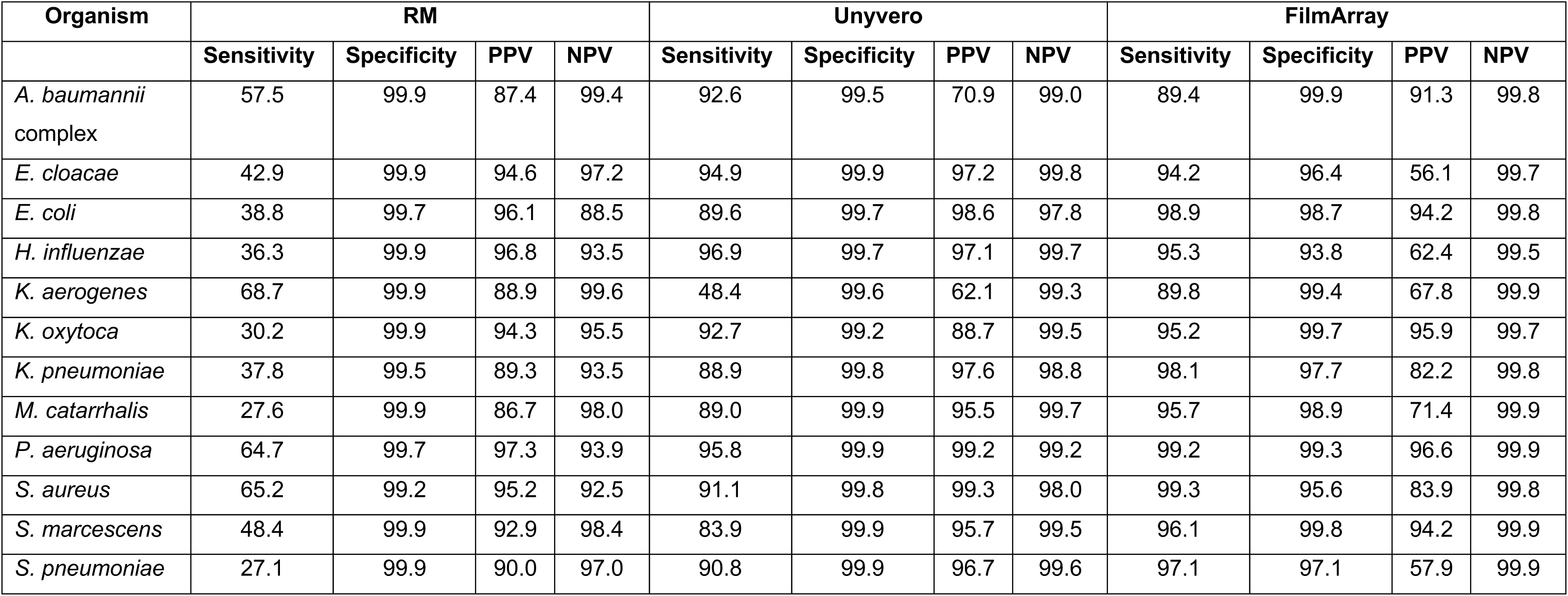
Pathogen-specific performance of RM and PCR tests estimated using BLC models. 95% credible intervals are omitted to aid readability but are shown in supplementary table S4. Only organisms on both PCR panels are included

Based on this BLC analysis, RM was the least sensitive technique for pathogen detection, with sensitivity values for individual pathogens ranging from 27.1 % for *Streptococcus pneumoniae* to 68.7% for *P. aeruginosa*. In contrast, sensitivity values for the PCR tests remained high; FilmArray sensitivity ranged from 89.4% to 99.3 %, whereas Unyvero sensitivity values ranged from 83.9% to 96.9% (expect *K. aerogenes*, 48.4%). Specificity and PPV values for both PCR tests increased considerably compared to the values calculated using RM as a gold standard: in particular, specificity was above 99% for Unyvero targets and ranged from 93.9% to 99.9% for FilmArray targets. The PPV range was 62.1% to 99.3% for Unyvero and 56.1% to 96.6% for FilmArray. These results are consistent with the observation that FilmArray detects pathogens more frequently than Unyvero.

### 16S Metagenomic Analysis

16S metagenomic analysis was originally included to act as an independent molecular reference method. Four-way BLC analysis including 16S metagenomic results is shown in Table 5. However, the 16S technique is only able to distinguish organisms to genus level, so PCR and RM data are likewise grouped to genus level. Streptococci are omitted because of the high density of commensal streptococci found in the respiratory tract and the inability of the 16S method to distinguish these from each other and from pathogenic species. For this analysis only, *Klebsiella aerogenes* was grouped within the genus *Enterobacter* owing to its relatively recent re-classification. The results show that 16S metagenomics was less sensitive than PCR and so was not an effective control method; nonetheless, it had had greater sensitivity compared with RM. Further optimisation may yield better results.

**Table 5.** Pathogen-specific performance of RM, PCR tests and 16S metagenomics estimated using BLC models. 95% credible intervals are omitted to aid readability but are shown in supplementary Table S5. Only organisms that can be detected by all four methods are included

| Organism | Routine Microbiology |  |  |  | Unyvero |  |  |  | FilmArray |  |  |  | 16S Metagenomics |  |  |  |
| --- | --- | --- | --- | --- | --- | --- | --- | --- | --- | --- | --- | --- | --- | --- | --- | --- |
|  | Sensitivity | Specificity | PPV | NPV | Sensitivity | Specificity | PPV | NPV | Sensitivity | Specificity | PPV | NPV | Sensitivity | Specificity | PPV | NPV |
| *Acinetobacter* | 54.2 | 99.9 | 87.5 | 99.2 | 92.9 | 99.7 | 83.0 | 99.9 | 83.2 | 99.9 | 91.4 | 99.7 | 83.2 | 97.8 | 38.9 | 99.7 |
| *Escherichia* | 38.7 | 99.6 | 95.3 | 88.3 | 95.6 | 99.8 | 99.2 | 99.1 | 99.2 | 97.8 | 90.9 | 99.8 | 73.3 | 99.2 | 94.9 | 94.5 |
| *Enterobacter* | 46.4 | 99.8 | 95.9 | 96.2 | 80.4 | 99.8 | 97.6 | 98.6 | 95.1 | 96.8 | 68.5 | 99.6 | 21.1 | 86.5 | 10.6 | 93.7 |
| *Haemophilus* | 30.4 | 99.8 | 96.6 | 90.9 | 80.5 | 99.7 | 97.1 | 97.3 | 96.8 | 95.5 | 75.8 | 99.5 | 85.2 | 88.0 | 50.5 | 97.6 |
| *Klebsiella* | 38.0 | 99.4 | 92.1 | 89.6 | 90.7 | 99.0 | 94.2 | 98.3 | 96.4 | 97.8 | 88.9 | 99.3 | 66.8 | 95.7 | 74.0 | 94.0 |
| *Moraxella* | 23.9 | 99.9 | 85.7 | 97.5 | 80.0 | 99.9 | 95.2 | 99.3 | 96.1 | 99.1 | 77.7 | 99.9 | 58.0 | 99.9 | 93.1 | 98.6 |
| *Pseudomonas* | 75.3 | 99.5 | 96.6 | 95.8 | 95.7 | 99.8 | 99.1 | 99.3 | 98.5 | 99.0 | 94.4 | 99.7 | 89.0 | 94.7 | 74.5 | 98.0 |
| *Proteus* | 33.6 | 99.7 | 84.8 | 96.5 | 96.6 | 99.7 | 94.7 | 99.8 | 96.9 | 99.3 | 88.9 | 99.8 | 54.6 | 99.9 | 95.5 | 97.6 |
| *Staphylococcus* | 66.3 | 99.6 | 97.5 | 92.5 | 92.6 | 99.8 | 99.2 | 98.2 | 99.3 | 95.6 | 84.5 | 99.8 | 82.8 | 77.9 | 47.4 | 95.0 |
| *Serratia* | 51.8 | 99.8 | 92.4 | 98.4 | 85.1 | 99.9 | 95.5 | 99.5 | 96.9 | 99.7 | 91.1 | 99.9 | 90.6 | 87.8 | 19.8 | 99.7 |

### Antimicrobial Resistance

All RM results for antimicrobial susceptibility testing were recorded and Table 5 shows rates of resistance to antimicrobials commonly used to treat HAP and VAP for the most-frequently-isolated species. *P. aeruginosa* exhibited the broadest resistance, with ≥ 10% rates for relevant antimicrobials. Other organisms, notably *E. coli* and *S. aureus*, had high rates of resistance to particular agents - 21.4% of *E. coli* were resistant to third-generation cephalosporins, and 18.4% of *S. aureus* were MRSA, however, all isolates of these species retained full susceptibility to other antimicrobials including meropenem for *E. coli* and glycopeptides for *S. aureus*. All species/genera had MDR rates of above 10%, with *E. coli* the highest at 37.1%.

**Table 5.** Antimicrobial resistance (%) to selected agents as determined by RM

|  | Antimicrobial |  |  |  |  |  |  |  |  |  |  |
| --- | --- | --- | --- | --- | --- | --- | --- | --- | --- | --- | --- |
| Organism | n<sup>a</sup> | AMC | 3GC | CIP | MAC | METH | GEN | MEM | TZP | GLYC | MDR |
| *P. aeruginosa* & *Pseudomonas* spp. | 79 | - | 21.6<sup>b</sup> | 17.3 | - | - | 10.8 | 23.6 | 24.3 | - | 20.3 |
| *S. aureus* | 83 | - | - | 18.9 | 25.0 | 14.6 | 10.0 | - | - | 0.0 | 13.3 |
| *E. coli* | 43 | 47.5 | 21.4 | 25.6 | - | - | 10.3 | 0.0 | 16.2 | - | 37.1 |
| *H. influenzae* | 23 | 22.2 | - | 0.0 | 41.7 | - | - | - | - | - | 13.0 |
| *Klebsiella* spp. | 44 | 30.8 | 20.7 | 10.5 | - | - | 7.7 | 3.3 | 13.5 | - | 15.9 |

- : Drug inherently inactive against species group
Antimicrobial abbreviations: AMC – amoxicillin/clavulanate, 3GC, third-generation cephalosporin (meaning ceftazidime, cefotaxime or ceftriaxone, as tested), CIP – ciprofloxacin, MAC – macrolide (erythromycin and clarithromycin depending on local laboratory), METH – detection of methicillin resistance (agent tested may be cefoxitin, flucloxacillin and oxacillin, depending on local laboratory); GEN – gentamicin, MEM – meropenem, TZP- piperacillin-tazobactam, GLYC, glycopeptide (vancomycin and teicoplanin depending on local laboratory)- MDR – multi-drug resistant, defined as resistant to $\geq 3$ classes according rules described in Magiorakos et al.<sup>23</sup>
<sup>a</sup> Refers to the total number of isolates in the data set. The number tested for any given drug may be fewer.
<sup>b</sup> Ceftazidime only considered for these species.

The PCR tests differ from RM by seeking resistance genes in a whole sample, rather than in particular bacteria. Very few ‘high concern’ resistance genes were found. Among carbapenemase genes the Unyvero found 6 acquired carbapenemase genes (2 *bla*_NDM_, 1 *bla*_KPC_ and 3 *bla*_VIM_) in addition to 5 *A. baumannii bla*_OXA-23_ genes, whereas FilmArray found 3 acquired carbapenemases (1 each of *bla*_KPC_, *bla*_IMP_ and *bla*_VIM_; it does not seen the genes encoding *Acinetobacter* OXA enzymes). Only two of these detections, one *bla*_KPC_ and one *bla*_VIM_ overlapped between the two tests. *bla*_CTX-M_, encoding the predominant family of ESBLs now seen in Enterobacterales was found by Unyvero in 14 specimens and by FilmArray in 32, with all Unyvero detections confirmed by the FilmArray. *mecA/C*, encoding methicillin resistance, was found in the presence of *S. aureus* by Unyvero in 25 specimens and by FilmArray in 32, with 18 of these in common between the two tests. Unyvero found 70 additional detections of *mecA/mecC* in the absence of *S. aureus*, this type of detection is prevented by design in the FilmArray, which amplifies across the junction between *mecA* and the *S. aureus* chromosome. Unyvero also seeks a wide range of resistance determinants, compromising unprotected penicillins, macrolides and sulphonamides; detections of these are summarised in Table S6; they were widely found but are more pertinent to antibiotics used in community pneumonia and were not considered further.

Assessment of the machines’ performance in respect of resistance gene detection is difficult because RM often reported no organisms for samples that were PCR positive for an organism and a resistance gene. In other cases, we were unable to retrieve routine clinical isolates for genetic validation. Where possible, RM isolates were independently investigated for the presence of antimicrobial resistance genes detected by the PCR tests. These isolates were supplemented with those recovered from our “comprehensive culture” methodology on a sub-set of discrepant samples; this aimed to recover all viable target species present, regardless of quantity. Following phenotypic antimicrobial susceptibility testing by disc diffusion, ESBL and carbapenemase genes were sought in Gram-negative bacteria using a Checkpoints microarray assay, whereas *mecA* and *mecC* genes were sought in *S. aureus* isolates by PCR. Concordance was then compared. Results are shown in table 6.

**Table 6.** Concordance of antimicrobial resistance gene detection by PCR and comparator methodology

| **Resistance Gene** | **Unyvero** |  |  |  | **FilmArray** |  |  |  |
| --- | --- | --- | --- | --- | --- | --- | --- | --- |
|  | Concordant detections<sup>a</sup>/total detections by PCR | Detections confirmed by RM | Additional detections confirmed by CC/number investigated<sup>b</sup> | Found in cultured isolates but missed in PCR testing | Concordant detections<sup>a</sup>/total detections by PCR | Detections confirmed by RM | Additional detections confirmed by CC/number investigated<sup>b</sup> | Found in cultured isolates but missed in PCR testing |
| *bla*<sub>CTX-M</sub> | 12/14 | 6 | 6/8 | 3 | 17/32 | 8 | 9/15 | 0 |
| *bla*<sub>IMP</sub> | 0/0 | 0 | 0/0 | 0 | 0/1 | 0 | 0/0 | 0 |
| *bla*<sub>KPC</sub> | 1/1 | 1 | 0/0 | 0 | 1/1 | 1 | 0/0 | 0 |
| *bla*<sub>OXA-23</sub> | 5/5 | 4 | 1/1 | 0 | NA | NA | NA | NA |
| *bla*<sub>OXA-24/40</sub>,<br>*bla*<sub>OXA-58</sub> | 0/0 | 0 | 0/0 | 0 | NA | NA | NA | NA |
| *bla*<sub>OXA-48</sub> | 0/0 | 0 | 0/0 | 0 | 0/0 | 0 | 0/0 | 0 |
| *bla*<sub>NDM</sub> | 0/2 | 0 | 0/2 | 0 | 0/0 | 0 | 0/0 | 0 |
| *bla*<sub>VIM</sub> | 2/3 | 2 | 0/1 | 0 | 1/1 | 0 | 0/0 | 1 |
| *mecA/mecC*<br>(+*MREJ* in FilmArray) | 13/25<sup>c</sup> | 10 | 3/5 | 1 | 15/32 | 12 | 3/8 | 0 |

<sup>a</sup>Total concordance from both RM and CC. Each sample is only counted once in the event of both tests being positive
<sup>b</sup>Excludes those where routine culture already found a positive isolate
<sup>c</sup>Only includes detections where *S. aureus* as well as *mecA/C* was also reported by the Unyvero. For total detections see table S5.

Among putative carbapenemase-producers, detections of *bla*_KPC_ (n=1) and OXA-23 (n= 5, Unyvero only) were in complete agreement, with the same genes detected in cultivated isolates. FilmArray missed 1/2 cases where the carbapenemase gene *bla*_VIM_ was detected in a cultivated isolated, and Unyvero missed 3/15 cases where the ESBL gene *bla*_CTX-M_ was present and 1/14 cases of MRSA. On the other hand, Unyvero detected one *bla*_VIM_ and two *bla*_NDM_ genes that could not be confirmed in cultivated isolates, FilmArray detected one *bla*_IMP_ that was not confirmed. Two *bla*_CTX-M_ detections by Unyvero, six *bla*_CTX-M_ detections by FilmArray, seven *mecA/C-*MREJ detections by Unyvero and six *mecA/C-*MREJ detections by FilmArray could not be confirmed by further culture-based investigation. Nine FilmArray detections of *bla*_CTX-M_ and 4 Unyvero, 11 FilmArray detections of *mecA/C-*MREJ were not investigated due to lack of materials and/or resources.

In total, comprehensive culture detected 12 additional organisms with confirmed resistance genes that were unrecorded by RM, which either had not isolated them or had discounted them.

### Overall Comparison of PCR Tests

Both PCR systems met the one essential requirement of having <5% major discordances. On this basis we collated all of the test results for both systems and combined them with performance and implementability data in order to choose which to carry forward to the INHALE RCT. Since, in this RCT, the machine would be located in the ICU rather than the microbiology laboratory and would require operation by medical and nursing staff, we sought to ensure that the chosen test was easy to utilise in this environment. Accordingly, our scoring scheme not only evaluated performance but also ease-of-use, footprint, turnaround time, as well as the overall user experience. This scheme was reviewed and agreed by INHALE’s independent Programme Steering Committee (PSC) as well as its Patient and Public Involvement (PPI) panel. The results are presented in Table 7.

**Table 7.** Scores allocated to PCR tests based on scoring system designed to evaluate overall performance, ease of use and implementability. See table S1 for full details of the scoring system.

| Criterion | Machine Score |  |  |  |
| --- | --- | --- | --- | --- |
|  | BioFire FilmArray Pneumonia Panel |  | Curetis Unyvero Pneumonia Panel |  |
|  | Value | Score | Value | Score |
| Overall concordance (max 45 points) | 71.3% | 16 | 74.8% | 20 |
| Sensitivity for detection of common pathogens (max 20 points) | 7 targets with better performance | 14 | 3 targets with better performance | 6 |
| Breadth of panel (max 15 points) | 191 unique detections | 12 | 244 unique detections | 15 |
| Time to result (max 15 points) | 75 min | 14 | 270 min | 7 |
| Cost per test (max 15 points)<sup>a</sup> | ++ | 15 | +++ | 10 |
| Failure rate (max 15 points) | 1.9% | 11 | 9.1%<sup>b</sup> | 0 |
| Footprint (max 5 points) | 3.2 sq. ft | 5 | 7.4 sq. ft | 1 |
| Customer service (max 5 points) | - | 4 | - | 3 |
| Consumable logistics (max 5 points)<sup>c</sup> | - | 5 | - | 0 |
| Ease of use (max 10 points) | - | 9 | - | 6 |
| **Total (Max 150)** | **-** | **105** | **-** | **68** |

<sup>a</sup> Costs in the range of £150-300/test depending on local purchase conditions. Includes estimates of cost of instrument purchase and operator time.
<sup>b</sup> includes both total and partial failures
<sup>c</sup>Comprised of one point each for space required for storage, storage temperature, delivery cost, delivery timescales and shelf-life.

FilmArray scored 105 points, compared with 68 for Unyvero. Overall, Unyvero was more concordant with RM but FilmArray had better sensitivity; Unyvero had a broader target panel but more failed tests. The FilmArray clearly performed better on characteristics relating to implementation, ease-of-use, turnaround time and user experience.

Accordingly, we have preferred the FilmArray Pneumonia Panel for the RCT, now being undertaken across 12 UK ICUs (Trial ID: ISRCTN16483855)

## Discussion

We have undertaken a comprehensive, manufacturer-independent, head-to-head comparison of the only two currently available rapid ‘sample-in, answer-out’ tests for the microbiological diagnosis of pneumonia; the Curetis Unyvero Hospitalized Pneumonia (HPN) Cartridge and the BioFire FilmArray Pneumonia Panel. All samples were collected from sick ICU patients for whom clinicians prescribed antimicrobials to treat pneumonia.

Both systems were considerably faster than RM and detected more pathogens. This reflected the poor sensitivity of RM in pneumonia.^9-11^ PCR tests tended to detect the *same* additional organisms in a given sample, over and above any reported by RM, implying that these additional detections were ‘*real’* and that PCR may improve microbiological diagnosis of pneumonia in ICU, increasing the proportion of patients swiftly receiving targeted antimicrobials. A confounder is that, unlike the molecular tests, RM was decentralised, being performed across 11 different hospital laboratories, receiving specimens from 15 ICUs. Notably, (and this will be discussed more extensively in a separate publication) RM showed considerable site-to site-variation, ranging from 29.5% to 85.7% sample positivity rate compared to 52.5 to 92.6% for PCR.

For analysis of test performance we initially chose RM, irrespective of source laboratory, as a gold standard. Whole-sample concordance analysis and individual pathogen-based sensitivity and specificity analyses were performed. Only 56.6 % of Unyvero results and 50.3 % of FilmArray results were fully concordant with RM, with the remaining partial concordances and minor discordances mostly reflecting additional organisms being detected by PCR. Per pathogen sensitivity performance was excellent for FilmArray, ranging from 91.7 to 100%; Unyvero performance was more variable, with sensitivity below 90% for several target pathogens. The numbers of cases where pathogens represented on the PCR panels were missed by these tests but found by culture were low. Both the sensitivity and specificity values are similar to those reported by others evaluating the same tests, as well as the regulatory clearance studies for the FilmArray Pneumonia Panel.^15,16,24-26^

We initially hoped that 16S metagenomics could act as an alternative, molecular, reference method; however, it proved less sensitive than PCR and this approach was abandoned. Instead, the limitations of RM, and the frequency with which both PCR tests detected the *same* organism that was missed by RM, led us to analyse the data also by BLC analysis. In simplified terms BLC analysis infers a new, unmeasurable yet present (i.e., latent) gold standard result. On the basis of this analysis, which makes no prior assumption about any one test being ‘correct’, (i) the sensitivity of RM appeared extremely poor, ranging from 27.1 to 68.7% according to the pathogen and (ii) both the specificity and PPV values for the PCR tests were considerably higher than those calculated using RM as the gold standard. Using this methodology, both PCR tests are clearly superior to RM. A caveat is that it is perhaps likely that two similar PCR tests (although they have different primers and different detection methods) should agree better with each other than with a dissimilar culture-based method. However, it is important to remember that all patients in this study were clinically diagnosed with respiratory infection and received contingent antibiotic treatment; it therefore seems more reasonable to consider an organism found by any one method as potentially significant rather than to dismiss the value of the method that most often recorded a potential pathogen.

A general concern about diagnostic PCR systems is that the additional organisms they find may prompt additional (and unnecessary) prescribing rather than good stewardship. Both systems offer semi-quantitative detection which may, in theory, assist with this issue. When we performed a sub-analysis, excluding organisms detected at low concentration by PCR, we did observe increased concordance with RM, however, there was also a substantial increase in missed detections. This suggests that improvements in concordance were mainly due to fewer organisms being detected by RM overall rather than any inherent propensity of RM to only detect and report bacteria at high concentrations. In large part, the issue with RM may reflect varying reporting practices at different RM laboratories. FilmArray’s manufacturer-led performance evaluation compared PCR detection with a reference-laboratory generated culture-based quantitative method and generally found good agreement between the two.^16^ Thus there may be scope for using the semi-quantitative results to inform therapy, particularly in cases where multiple organisms are present. This approach would require thorough validation by comparison with clinical indicators.

The types and relative frequencies of organisms identified in these HAP and VAP patients were similar for RM and both PCR tests, without any obvious bias for culture (or PCR) to miss particular organisms. The species distribution resembled that reported in numerous HAP/VAP studies from Europe and North America, with *S. aureus, P. aeruginosa* and Enterobacterales the predominant organisms^7,8^. Collated antimicrobial susceptibility data generated by the RM laboratories revealed that the prevalence of antimicrobial resistance was generally high. In particular, both *P. aeruginosa* and *S. aureus* isolates had higher rates of resistance to the antibiotics used as standard-of-care in HAP/VAP than reported among nationally-collected LRTI isolates in the BSAC national surveillance programme (2017/18 data). Higher rates of resistance may reflect the nature of ICU HAP/VAP patients, who frequently receive multiple courses of broad-spectrum antimicrobials; by contrast, only around half the BSAC-tested HAP/VAP isolates are from ICU patients, with the remainder originating from patients on general wards.

Comparison of antimicrobial resistance gene detection by FilmArray and Unyvero with RM antimicrobial susceptibility testing data is complicated by the imperfect nature of genotype / phenotype associations: genes may be carried but not expressed, and phenotypic resistance may arise from mechanisms that were not sought (for example a combination of an ESBL and impermeability may confer carbapenem resistance).^27^ Moreover, direct detection of a resistance gene in a clinical sample does not indicate which bacterial species is hosting that gene. We therefore conducted our own independent genotypic verification of bacterial isolates identified as phenotypically resistant by RM and for further organisms recovered by Comprehensive Culture, which sought to recover isolates not retained or considered significant by the routine laboratories. Overall, 65% of Unyvero detections and 50% of FilmArray detections were concordant against a combination of RM and CC results. Of the remainder, the majority of discordance related to cases where it was not possible to perform CC, although some genuine discordances were identified. Crucially, the PCR tests identified several key high-consequence resistance genes, later confirmed by CC to be present in viable bacteria, that had been missed by RM. Although rapid PCR-based detection of resistance genes does not provide the user with a full susceptibility profile, it is a swift and sensitive predictor of resistance and might be useful for early identification of patients who need to be isolated or to have their therapy escalated. A complication, in the case of *S. aureus*/MRSA is that the Unyvero Pneumonia Panel reports all *mecA* and *mecC* detections regardless of whether *S. aureus* is found, resulting in many detections that may reflect the presence of *mecA* in coagulase-negative staphylococci (Table S6); this is not an issue for FilmArray, which specifically seeks *mecA/C* in *S. aureus*.^*28*^ In our concordance analysis we only considered *mecA and mecC* when reported together with *S. aureus*.

Unyvero seeks a range of further resistance determinants (*ermB, bla*_TEM_ *bla*_SHV_, *sul1* and *gyrA* mutations) but these are of lower relevance to the antibiotics likely to be used in HAP, being more pertinent when the test is used for CAP pathogens. Moreover, most are very difficult to associate with a particular organism owing to their wide distribution in commensal flora. Both *ermB* and *bla*_TEM_ were widely found by nanopore sequencing in respiratory samples of individuals harbouring only ‘normal flora’.^29^ Accordingly we chose not to verify them in the present study.

The run times of the machines are measured in hours rather than the days required for standard of care culture. However, total turn-around will also depend on the machine’s placement in the clinical pathway; this could not be measured here because tests were run retrospectively under research conditions, not in clinical settings. However, we established that the median transport time of samples from the ICU to the laboratory was 6h, with longer times when laboratories were remote from the hospital site; this interval is around the same period required for the Unyvero test and 4 times longer than for the FilmArray’s test. If the advantages of speed are to be realised, it follows that the machine must be place in, or near to, the ICU itself.

The decision of whether or not to adopt a rapid diagnostic test into routine clinical practice will depend not only on its performance but also on the practicalities of implementation. Here, we evaluated diagnostic accuracy as well as potential for implementation, and on that basis, found the FilmArray to be not only more sensitive than the Unyvero but also faster, smaller and easier to use. Accordingly, we have chosen the FilmArray Pneumonia panel to take forward into the next phase of our research, involving a trial where patients are randomised to treatment guided by the test results or to ‘standard to care’, comprising empirical antibiotics, adapted once microbiology results become available. This trial is now in progress across 12 UK ICUs and will evaluate whether the test improves antimicrobial stewardship and patient outcomes. So that the advantages of speed can be realised, the machines have been placed in the ICUs themselves, not in microbiology laboratories, and are run by ICU staff. Their gene and species outputs are interpreted using a prescribing algorithm, co-developed with local microbiologists, who will also review treatments given in their (typically daily) ICU ward rounds.

The increased sensitivity and speed of these PCR tests compared with culture warrants their further evaluation for potential adoption into clinical practice. Currently, the poor performance of RM is tolerated and the majority of antimicrobial prescribing for HAP and VAP remains empirical, utilising broad-spectrum antimicrobials for prolonged periods, despite this being sub-optimal stewardship. Rapid diagnostics could improve patient outcomes and support early use of targeted antimicrobials. This approach may be especially beneficial for example against *S. aureus*, where several targeted, narrow-spectrum, antimicrobial options are available and are likely to cause less disturbance of the gut flora than prolonged use of broad-spectrum agents. Bolder deployment of sensitive PCR diagnosis may also involve de-escalating or stopping antimicrobials in cases where no pathogens are detected.

In summary, faster and more sensitive PCR tests for the diagnosis of severe lower respiratory tract offer considerable potential for improved antimicrobial stewardship and more targeted and personalised treatment of pneumonia; RCT data are eagerly awaited to determine if this potential can be realised without compromising patient safety.^30^

## Data Availability

Data available upon request

## Acknowledgements

We would like to offer our sincerest gratitude to all ICU and laboratory staff at participating sites who assisted with sample identification and collection. We would also like to thank Norwich Clinical Trials Unit and the INHALE PPI group for their support.

## Transparency declarations

### DML

Advisory Boards or ad-hoc consultancy Accelerate, Allecra, Antabio, Centauri, Entasis, GSK, Meiji, Melinta, Menarini, Mutabilis, Nordic, ParaPharm, Pfizer, QPEX, Roche, Shionogi, T.A.Z., Tetraphase, VenatoRx, Wockhardt, Zambon, Paid lectures – Astellas, bioMérieux, Beckman Coulter, Cardiome, Cepheid, Merck/MSD, Menarini, Nordic, Pfizer and Shionogi. Relevant shareholdings or options – Dechra, GSK, Merck, Perkin Elmer, Pfizer, T.A.Z, amounting to <10% of portfolio value.

### VG

Advisory boards or ad-hoc consultancy Gilead, Shionogi, bioMérieux, MSD, Vidya Diagnostics

### VE

Speaking honoraria, consultancy fees and in-kind contributions from several diagnostic companies including Curetis GmbH, bioMérieux and Oxford Nanopore.

### JOG

JOG: has received speaking honoraria, consultancy fees, in-kind contributions or research funding from Oxford Nanopore, Simcere, Becton-Dickinson and Heraeus Medical.

### All other authors

None to declare.

