## Supplementary data for "Multicentre evaluation of two multiplex PCR platforms for the rapid microbiological investigation of nosocomial pneumonia in UK ICUs: the INHALE WP1 study"

**Table S1.** Criteria for HAP/VAP diagnostic test progression to RCT phase of the inhale study (WP3)

| **Criterion** | **Description** | **Point Scoring** | **Maximum available points** |
|---|---|---|---|
| Concordance - essential criterion | Major discordance i.e. failures by the test to find pathogen(s) detected by routine microbiology must account for < 5% of all tests performed. | NA | NA |
| Overall Concordance | A measure of the overall accuracy of the test compared to the gold standard. | 1 point is awarded for every % point over 55% overall concordance | 45 points |
| Sensitivity | Sensitivity for detection of common pathogens (i.e. *P. aeruginosa, S. aureus, K. pneumoniae, K. oxytoca*, *E. coli, E. cloacae, E. aerogenes, A. baumannii, H. influenzae* and *S. pneumoniae*) | 2 points for every 'win', i.e. the best sensitivity against a particular pathogen | 20 points |
| Breadth of panel | Each PCR test seeks some targets that the other cannot, principally resistance genes for Curetis and viruses for Biofire. | Maximum points for most detections of unique targets, other tests awarded points as a proportion of unique detection | 15 points |
| Time to Result | Time to Result | 1 Point allocated for each 30 min less than 8h, the common dosage interval for antibiotics | 15 points |
| Cost of tests and equipment | Cost per test, A composite measure of both test and equipment cost. | Cheapest test is awarded the maximum points. One point is deducted from others for every 10% increase in price compared to the cheapest. | 15 points |
| Failure rate | Failure rate of test and/or machine, full or partial. | 1 point deducted for each 0.5% of failures | 15 points |
| Footprint and space occupied | Amount of space required to host machine | Smallest machine awarded maximum points. Cheapest test is awarded the maximum points. One point is deducted from others for every 10% increase in price compared to the cheapest. | 5 points |
| Customer service | The quality and speed of customer service in the event of breakdown, ordering, installation etc. | . Average score based on assessment from individual users who have dealt with manufacturers during the study. | 5 points |
| Consumable logistics | Space required for storage of consumables, storage temperature, shelf life, delivery speed, delivery cost. | 1 point for best performing machine for each criterion | 5 points |
| Ease of use | User perception and experience | Average scored based on assessments from individual users who have operated machines during the study. | 10 points |
| **Total** | | | **150 points** |

**Table S2**. Viral detections made by FilmArray (n=620 eligible samples) and sensitivity and specificity compared to routine virology. 95% confidence intervals are given in brackets.

| **Virus** | **Number of Detections** | **% positive samples** | **Sensitivity %** | **Specificity %** | **PPV %** | **NPV %** |
|---|---|---|---|---|---|---|
| Rhinovirus | 55 | 8.9 | 100.0 (15.8-100.0) | 91.4 (88.9-93.5) | 3.6 (0.4-12.5) | 100.0 (99.3-100.0) |
| Influenza A | 29 | 4.7 | 100.0 (59.0-100.0) | 96.4 (94.6-97.7) | 24.1 (10.3-43.5) | 100.0 (99.4-100.0) |
| Influenza B | 25 | 4.0 | 75.0 (19.4-99.4) | 96.6 (94.8-97.9) | 16.0 (4.5-36.1) | 100.0 (99.4-100.0) |
| Parainfluenza | 17 | 2.7 | 75.0 (19.4-99.4) | 97.7 (96.2-98.8) | 17.6 (3.8-43.4) | 99.8 (99.1-100.0) |
| Coronavirus (229E, HKU1, NL63, OC43) | 16 | 2.6 | ND* | 97.4 (95.8-98.5) | 0.0 (0.0-20.6) | 100.0 (99.4-100.0) |
| Adenovirus | 7 | 1.1 | 50.0 (6.8-93.2) | 99.2 (98.1-99.7) | 28.6 (3.7-71.0) | 99.7 (98.8-100.0) |
| Respiratory Syncytial Virus | 6 | 1.0 | 66.7 (9.4-99.2) | 99.4 (98.3-99.8) | 33.3 (4.3-77.7) | 99.8 (99.1-100.0) |
| Human metapneumovirus | 5 | 0.8 | 100.0 (2.5-100.0) | 99.4 (98.4-99.8) | 20.0 (0.5-71.6) | 100.0 (99.4-100.0) |
| MERS coronavirus | 0 | 0 | ND | ND | ND | ND |

*ND – not determined because routine virology did not report any positives

**Table S3.** Pathogen specific performance of PCR tests when compared to routine microbiology as the gold standard including 95% confidence intervals

| UNYVERO | | | | | | | | | | | | | |
|---|---|---|---|---|---|---|---|---|---|---|---|---|---|
| **Target Organism** | **Number of Detections** | **Sensitivity** | | | **Specificity** | | | **PPV** | | | **NPV** | | |
| | | **%** | **95% CI** | | **%** | **95% CI** | | **%** | **95% CI** | | **%** | **95% CI** | |
| *P. aeruginosa* | 94 | 95.3 | 86.9 | 99.0 | 93.9 | 91.6 | 95.8 | 64.9 | 54.4 | 74.5 | 99.4 | 98.3 | 99.9 |
| *S. aureus* | 104 | 87.2 | 77.7 | 93.7 | 93.2 | 90.7 | 95.2 | 65.4 | 55.4 | 74.4 | 98.0 | 96.4 | 99.0 |
| *K. pneumoniae* | 54 | 83.3 | 62.6 | 95.3 | 94.2 | 91.9 | 95.9 | 37.0 | 24.3 | 51.3 | 99.3 | 98.2 | 99.8 |
| *K. oxytoca* | 40 | 90.9 | 58.7 | 99.8 | 95.0 | 92.9 | 96.6 | 25.0 | 12.7 | 41.2 | 99.8 | 99.0 | 100.0 |
| *E. coli* | 96 | 87.8 | 73.8 | 95.9 | 89.4 | 86.5 | 91.8 | 37.5 | 27.8 | 48.0 | 99.0 | 97.7 | 99.7 |
| *E. cloacae* | 27 | 100.0 | 73.5 | 100.0 | 97.5 | 95.9 | 98.6 | 44.4 | 25.5 | 64.7 | 100.0 | 99.4 | 100.0 |
| *K. aerogenes* | 6 | 50.0 | 11.8 | 88.2 | 99.5 | 98.5 | 99.9 | 50.0 | 11.8 | 88.2 | 99.5 | 98.5 | 99.9 |
| *A. baumannii* | 11 | 100.0 | 47.8 | 100.0 | 99.0 | 97.8 | 99.6 | 45.5 | 16.7 | 76.6 | 100.0 | 99.4 | 100.0 |
| *H. influenzae* | 58 | 100.0 | 83.9 | 100.0 | 93.7 | 91.4 | 95.5 | 36.2 | 24.0 | 49.9 | 100.0 | 99.3 | 100.0 |
| *S. pneumoniae* | 22 | 100.0 | 54.1 | 100.0 | 97.3 | 95.7 | 98.5 | 27.3 | 10.7 | 50.2 | 100.0 | 99.4 | 100.0 |
| *M. catarrhalis* | 15 | 100.0 | 39.8 | 100.0 | 98.2 | 96.8 | 99.1 | 26.7 | 7.8 | 55.1 | 100.0 | 99.4 | 100.0 |
| *S. marcescens* | 17 | 77.8 | 40.0 | 97.2 | 98.3 | 96.9 | 99.2 | 41.2 | 18.4 | 67.1 | 99.7 | 98.8 | 100.0 |
| *C. pneumoniae* | 0 | | | | | | | | | | | | |
| *L. pneumophila* | 2 | | | | | | | | | | | | |
| *M. pneumoniae* | 1 | | | | | | | | | | | | |
| *C. freundii* | 9 | 100.0 | 2.5 | 100.0 | 98.7 | 97.4 | 99.4 | 11.1 | 0.3 | 48.2 | 100.0 | 99.4 | 100.0 |
| *M. morganii* | 11 | 100.0 | 2.5 | 100.0 | 98.3 | 97.0 | 99.2 | 9.1 | 0.2 | 41.3 | 100.0 | 99.4 | 100.0 |
| *S. maltophilia* | 46 | 92.9 | 66.1 | 99.8 | 94.4 | 92.3 | 96.1 | 28.3 | 16.0 | 43.5 | 99.8 | 99.0 | 100.0 |

| FILMARRAY | | | | | | | | | | | | | |
|---|---|---|---|---|---|---|---|---|---|---|---|---|---|
| **Target Organism** | **Number of Detections** | Sensitivity | | | Specificity | | | PPV | | | NPV | | |
| | | % | 95% CI | | % | 95% CI | | % | 95% CI | | % | 95% CI | |
| *P. aeruginosa* | 103 | 98.5 | 91.8 | 100.0 | 93.1 | 90.7 | 95.1 | 63.1 | 53.0 | 72.4 | 99.8 | 98.9 | 100.0 |
| *S. aureus* | 137 | 96.2 | 89.4 | 99.2 | 88.9 | 85.9 | 91.4 | 56.2 | 47.5 | 64.7 | 99.4 | 98.2 | 99.9 |
| *K. pneumoniae* | 74 | 92.0 | 74.0 | 99.0 | 91.4 | 88.9 | 93.6 | 31.1 | 20.8 | 42.9 | 99.6 | 98.7 | 100.0 |
| *K. oxytoca* | 40 | 100.0 | 71.5 | 100.0 | 95.2 | 93.2 | 96.8 | 27.5 | 14.6 | 43.9 | 100.0 | 99.4 | 100.0 |
| *E. coli* | 113 | 97.6 | 87.4 | 99.9 | 87.5 | 84.6 | 90.1 | 36.3 | 27.4 | 45.9 | 99.8 | 98.9 | 100.0 |
| *E. cloacae* | 51 | 91.7 | 61.5 | 99.8 | 93.4 | 91.1 | 95.3 | 21.6 | 11.3 | 35.3 | 99.8 | 99.0 | 100.0 |
| *K. aerogenes* | 11 | 100.0 | 54.1 | 100.0 | 99.2 | 98.1 | 99.7 | 54.5 | 23.4 | 83.3 | 100.0 | 99.4 | 100.0 |
| *A. baumannii* | 9 | 100.0 | 54.1 | 100.0 | 99.5 | 98.6 | 99.9 | 66.7 | 29.9 | 92.5 | 100.0 | 99.4 | 100.0 |
| *H. influenzae* | 91 | 95.2 | 76.2 | 99.9 | 88.1 | 85.3 | 90.6 | 22.0 | 14.0 | 31.9 | 99.8 | 99.0 | 100.0 |
| *S. pneumoniae* | 40 | 100.0 | 54.1 | 100.0 | 94.5 | 92.3 | 96.1 | 15.0 | 5.7 | 29.8 | 100.0 | 99.4 | 100.0 |
| *M. catarrhalis* | 23 | 100.0 | 39.8 | 100.0 | 96.9 | 95.2 | 98.1 | 17.4 | 5.0 | 38.8 | 100.0 | 99.4 | 100.0 |
| *S. marcescens* | 20 | 100.0 | 66.4 | 100.0 | 98.2 | 96.8 | 99.1 | 45.0 | 23.1 | 68.5 | 100.0 | 99.4 | 100.0 |
| *C. pneumoniae* | 0 | | | | | | | | | | | | |
| *L. pneumophila* | 0 | | | | | | | | | | | | |
| *M. pneumoniae* | 1 | | | | | | | | | | | | |
| *S. agalactiae* | 22 | NA | 0.0 | 100.0 | 96.5 | 94.7 | 97.8 | 0.0 | 0.0 | 15.4 | 100.0 | 99.4 | 100.0 |
| *S. pyogenes* | 9 | 100.0 | 15.8 | 100.0 | 98.9 | 97.7 | 99.5 | 22.2 | 2.8 | 60.0 | 100.0 | 99.4 | 100.0 |

**Table S4.** Pathogen specific performance of routine microbiology and PCR tests using independent BLC modelling including 95% confidence intervals

| ROUTINE MICROBIOLOGY | | | | | | | | | | | | |
|---|---|---|---|---|---|---|---|---|---|---|---|---|
| **Target Organism** | **Sensitivity** | | | **Specificity** | | | **PPV** | | | **NPV** | | |
| | **%** | **95% CI** | | **%** | **95% CI** | | **%** | **95% CI** | | **%** | **95% CI** | |
| *P. aeruginosa* | 64.7 | 54.7 | 73.9 | 99.7 | 98.9 | 100.0 | 97.3 | 91.5 | 99.6 | 93.9 | 91.7 | 95.8 |
| *S. aureus* | 65.2 | 56.1 | 74.1 | 99.2 | 98.2 | 99.8 | 95.2 | 88.8 | 98.6 | 92.5 | 90.0 | 94.7 |
| *K. pneumoniae* | 37.8 | 26.0 | 51.4 | 99.5 | 98.6 | 99.9 | 89.3 | 73.2 | 97.7 | 93.5 | 90.9 | 95.6 |
| *K. oxytoca* | 30.2 | 18.3 | 45.5 | 99.9 | 99.3 | 100.0 | 94.3 | 73.0 | 99.8 | 95.5 | 93.7 | 97.0 |
| *E. coli* | 38.8 | 29.8 | 48.2 | 99.7 | 98.9 | 100.0 | 96.1 | 86.8 | 99.5 | 88.5 | 85.5 | 91.1 |
| *E. cloacae* | 42.9 | 25.6 | 61.3 | 99.9 | 99.3 | 100.0 | 94.6 | 71.6 | 99.8 | 97.2 | 95.5 | 98.4 |
| *K. aerogenes* | 68.7 | 32.1 | 94.7 | 99.9 | 99.4 | 100.0 | 88.9 | 54.4 | 99.6 | 99.6 | 98.5 | 99.9 |
| *A. baumannii* | 57.5 | 27.1 | 84.9 | 99.9 | 99.4 | 100.0 | 87.4 | 50.8 | 99.5 | 99.4 | 98.5 | 99.8 |
| *H. influenzae* | 36.3 | 24.8 | 49.1 | 99.9 | 99.3 | 100.0 | 96.8 | 84.5 | 99.9 | 93.5 | 91.0 | 95.4 |
| *S. pneumoniae* | 27.1 | 15.9 | 46.2 | 99.9 | 99.4 | 100.0 | 90.0 | 61.0 | 99.6 | 97.0 | 95.0 | 98.3 |
| *M. catarrhalis* | 27.6 | 15.8 | 50.4 | 99.9 | 99.4 | 100.0 | 86.7 | 50.5 | 99.4 | 98.0 | 96.5 | 99.0 |
| *S. marcescens* | 48.4 | 27.7 | 69.7 | 99.9 | 99.3 | 100.0 | 92.9 | 67.4 | 99.7 | 98.4 | 97.1 | 99.2 |

| UNYVERO | | | | | | | | | | | | |
|---|---|---|---|---|---|---|---|---|---|---|---|---|
| **Target Organism** | **Sensitivity** | | | **Specificity** | | | **PPV** | | | **NPV** | | |
| | **%** | **95% CI** | | **%** | **95% CI** | | **%** | **95% CI** | | **%** | **95% CI** | |
| *P. aeruginosa* | 95.8 | 89.6 | 99.0 | 99.9 | 99.2 | 100.0 | 99.2 | 95.8 | 100.0 | 99.2 | 98.0 | 99.8 |
| *S. aureus* | 91.1 | 82.9 | 96.1 | 99.8 | 99.2 | 100.0 | 99.3 | 96.4 | 100.0 | 98.0 | 95.9 | 99.2 |
| *K. pneumoniae* | 88.9 | 73.3 | 97.5 | 99.8 | 99.0 | 100.0 | 97.6 | 90.6 | 99.9 | 98.8 | 96.6 | 99.8 |
| *K. oxytoca* | 92.7 | 80.3 | 98.8 | 99.2 | 98.1 | 99.9 | 88.7 | 74.7 | 98.6 | 99.5 | 98.6 | 99.9 |
| *E. coli* | 89.6 | 80.5 | 96.4 | 99.7 | 98.9 | 100.0 | 98.6 | 94.4 | 99.9 | 97.8 | 95.8 | 99.3 |

| | | | | | | | | | | | | |
|---|---|---|---|---|---|---|---|---|---|---|---|---|
| *E. cloacae* | 94.9 | 74.9 | 99.8 | 99.9 | 99.3 | 100.0 | 97.2 | 86.9 | 99.9 | 99.8 | 98.5 | 100.0 |
| *K. aerogenes* | 48.4 | 21.4 | 80.3 | 99.6 | 98.8 | 99.9 | 62.1 | 25.5 | 93.0 | 99.3 | 98.1 | 99.8 |
| *A. baumannii* | 92.6 | 66.2 | 99.7 | 99.5 | 98.6 | 99.9 | 70.9 | 39.7 | 94.6 | 99.9 | 99.4 | 100.0 |
| *H. influenzae* | 96.9 | 84.8 | 99.9 | 99.7 | 98.8 | 100.0 | 97.1 | 89.3 | 99.9 | 99.7 | 98.2 | 100.0 |
| *S. pneumoniae* | 90.8 | 63.2 | 99.6 | 99.9 | 99.3 | 100.0 | 96.7 | 83.8 | 99.9 | 99.6 | 97.8 | 100.0 |
| *M. catarrhalis* | 89.0 | 60.6 | 99.5 | 99.9 | 99.4 | 100.0 | 95.5 | 78.1 | 99.8 | 99.7 | 98.5 | 100.0 |
| *S. marcescens* | 83.9 | 64.1 | 95.6 | 99.9 | 99.4 | 100.0 | 95.7 | 78.0 | 99.8 | 99.5 | 98.7 | 99.9 |

| **FILMARRAY** | | | | | | | | | | | | |
|---|---|---|---|---|---|---|---|---|---|---|---|---|
| | **Sensitivity** | | | **Specificity** | | | **PPV** | | | **NPV** | | |
| **Target Organism** | **%** | **95% CI** | | **%** | **95% CI** | | **%** | **95% CI** | | **%** | **95% CI** | |
| *P. aeruginosa* | 99.2 | 95.9 | 100.0 | 99.3 | 98.3 | 99.9 | 96.6 | 91.1 | 99.6 | 99.9 | 99.2 | 100.0 |
| *S. aureus* | 99.3 | 96.5 | 100.0 | 95.6 | 93.3 | 97.5 | 83.9 | 76.2 | 90.7 | 99.8 | 99.1 | 100.0 |
| *K. pneumoniae* | 98.1 | 91.1 | 99.9 | 97.7 | 95.8 | 99.4 | 82.2 | 69.5 | 95.6 | 99.8 | 99.0 | 100.0 |
| *K. oxytoca* | 95.2 | 81.0 | 99.8 | 99.7 | 98.9 | 100.0 | 95.9 | 84.0 | 99.8 | 99.7 | 98.6 | 100.0 |
| *E. coli* | 98.9 | 95.3 | 100.0 | 98.7 | 96.8 | 99.9 | 94.2 | 86.1 | 99.6 | 99.8 | 99.0 | 100.0 |
| *E. cloacae* | 94.2 | 81.7 | 99.2 | 96.4 | 94.6 | 97.9 | 56.1 | 40.5 | 72.6 | 99.7 | 99.0 | 100.0 |
| *K. aerogenes* | 89.8 | 58.5 | 99.6 | 99.4 | 98.4 | 99.9 | 67.8 | 34.3 | 96.7 | 99.9 | 99.2 | 100.0 |
| *A. baumannii* | 89.4 | 55.6 | 99.5 | 99.9 | 99.4 | 100.0 | 91.3 | 62.5 | 99.6 | 99.8 | 99.2 | 100.0 |
| *H. influenzae* | 95.3 | 87.4 | 99.2 | 93.8 | 91.5 | 95.8 | 62.4 | 51.9 | 73.8 | 99.5 | 98.5 | 99.9 |
| *S. pneumoniae* | 97.1 | 85.8 | 99.9 | 97.1 | 95.4 | 98.8 | 57.9 | 40.8 | 81.7 | 99.9 | 99.3 | 100.0 |
| *M. catarrhalis* | 95.7 | 80.2 | 99.8 | 98.9 | 97.6 | 99.8 | 71.4 | 47.1 | 95.0 | 99.9 | 99.4 | 100.0 |
| *S. marcescens* | 96.1 | 81.8 | 99.9 | 99.8 | 99.2 | 100.0 | 94.2 | 76.3 | 99.8 | 99.9 | 99.3 | 100.0 |

**Table S5.** Pathogen-specific performance of routine microbiology, PCR tests and 16S metagenomics using independent BLC modelling, showing 95% confidence intervals

| ROUTINE MICROBIOLOGY | | | | | | | | | | | | |
|---|---|---|---|---|---|---|---|---|---|---|---|---|
| | Sensitivity | | | Specificity | | | PPV | | | NPV | | |
| **Target Genus** | **%** | **95% CI** | | **%** | **95% CI** | | **%** | **95% CI** | | **%** | **95% CI** | |
| *Acinetobacter* | 54.2 | 26.8 | 82.0 | 99.9 | 99.3 | 100.0 | 87.5 | 49.5 | 99.5 | 99.2 | 98.3 | 99.8 |
| *Escherichia* | 38.7 | 29.1 | 48.7 | 99.6 | 98.6 | 99.9 | 95.3 | 85.2 | 99.3 | 88.3 | 85.2 | 91.0 |
| *Enterobacter* | 46.4 | 30.2 | 63.3 | 99.8 | 99.2 | 100.0 | 95.9 | 79.0 | 99.9 | 96.2 | 93.7 | 97.8 |
| *Haemophilus* | 30.4 | 20.2 | 42.8 | 99.8 | 99.2 | 100.0 | 96.6 | 83.7 | 99.9 | 90.9 | 87.7 | 93.4 |
| *Klebsiella* | 38.0 | 27.8 | 48.5 | 99.4 | 98.3 | 99.8 | 92.1 | 79.4 | 98.1 | 89.6 | 86.6 | 92.3 |
| *Moraxella* | 23.9 | 15.4 | 43.4 | 99.9 | 99.3 | 100.0 | 85.7 | 50.4 | 99.5 | 97.5 | 96.0 | 98.6 |
| *Pseudomonas* | 75.3 | 64.8 | 83.8 | 99.5 | 98.5 | 100.0 | 96.6 | 89.8 | 99.8 | 95.8 | 93.7 | 97.4 |
| *Proteus* | 33.6 | 18.3 | 52.2 | 99.7 | 98.9 | 100.0 | 84.8 | 57.4 | 97.7 | 96.5 | 94.6 | 97.8 |
| *Staphylococcus* | 66.3 | 56.4 | 75.2 | 99.6 | 98.6 | 99.9 | 97.5 | 91.6 | 99.7 | 92.5 | 89.6 | 94.8 |
| *Serratia* | 51.8 | 30.3 | 73.7 | 99.8 | 99.2 | 100.0 | 92.4 | 65.7 | 99.7 | 98.4 | 97.1 | 99.3 |

| UNYVERO | | | | | | | | | | | | |
|---|---|---|---|---|---|---|---|---|---|---|---|---|
| | Sensitivity | | | Specificity | | | PPV | | | NPV | | |
| **Target Genus** | **%** | **95% CI** | | **%** | **95% CI** | | **%** | **95% CI** | | **%** | **95% CI** | |
| *Acinetobacter* | 92.9 | 67.9 | 99.7 | 99.7 | 98.8 | 100.0 | 83.0 | 51.9 | 98.0 | 99.9 | 99.3 | 100.0 |
| *Escherichia* | 95.6 | 89.4 | 99.0 | 99.8 | 99.1 | 100.0 | 99.2 | 96.0 | 100.0 | 99.1 | 97.7 | 99.8 |
| *Enterobacter* | 80.4 | 60.6 | 94.3 | 99.8 | 99.2 | 100.0 | 97.6 | 87.7 | 99.9 | 98.6 | 96.4 | 99.6 |
| *Haemophilus* | 80.5 | 67.5 | 92.0 | 99.7 | 98.8 | 100.0 | 97.1 | 90.6 | 99.8 | 97.3 | 95.0 | 99.0 |
| *Klebsiella* | 90.7 | 81.8 | 96.6 | 99.0 | 97.6 | 99.8 | 94.2 | 86.5 | 98.8 | 98.3 | 96.5 | 99.4 |
| *Moraxella* | 80.0 | 56.0 | 95.3 | 99.9 | 99.3 | 100.0 | 95.2 | 76.8 | 99.9 | 99.3 | 98.2 | 99.9 |
| *Pseudomonas* | 95.7 | 89.4 | 99.0 | 99.8 | 99.2 | 100.0 | 99.1 | 95.2 | 100.0 | 99.3 | 98.1 | 99.8 |
| *Proteus* | 96.6 | 83.5 | 99.8 | 99.7 | 99.0 | 100.0 | 94.7 | 82.5 | 99.7 | 99.8 | 99.0 | 100.0 |
| *Staphylococcus* | 92.6 | 85.2 | 97.1 | 99.8 | 99.1 | 100.0 | 99.2 | 96.1 | 100.0 | 98.2 | 96.3 | 99.3 |
| *Serratia* | 85.1 | 64.5 | 96.3 | 99.9 | 99.3 | 100.0 | 95.5 | 78.0 | 99.8 | 99.5 | 98.7 | 99.9 |

| FILMARRAY | | | | | | | | |
|---|---|---|---|---|---|---|---|---|
| | Sensitivity | | Specificity | | PPV | | NPV | |
| **Target Genus** | **%** | **95% CI** | **%** | **95% CI** | **%** | **95% CI** | **%** | **95% CI** |

| | | | | | | | | | | | | |
|---|---|---|---|---|---|---|---|---|---|---|---|---|
| *Acinetobacter* | 83.2 | 53.5 | 98.4 | 99.9 | 99.2 | 100.0 | 91.4 | 61.8 | 99.7 | 99.7 | 99.0 | 100.0 |
| *Escherichia* | 99.2 | 95.9 | 100.0 | 97.8 | 96.0 | 99.0 | 90.9 | 83.7 | 95.9 | 99.8 | 99.1 | 100.0 |
| *Enterobacter* | 95.1 | 84.6 | 99.4 | 96.8 | 94.6 | 98.7 | 68.5 | 51.9 | 86.8 | 99.6 | 98.8 | 100.0 |
| *Haemophilus* | 96.8 | 89.8 | 99.5 | 95.5 | 92.9 | 97.6 | 75.8 | 63.2 | 86.7 | 99.5 | 98.5 | 99.9 |
| *Klebsiella* | 96.4 | 90.0 | 99.7 | 97.8 | 95.9 | 99.1 | 88.9 | 79.7 | 95.8 | 99.3 | 98.0 | 99.9 |
| *Moraxella* | 96.1 | 81.2 | 99.9 | 99.1 | 97.9 | 99.8 | 77.7 | 54.3 | 95.8 | 99.9 | 99.3 | 100.0 |
| *Pseudomonas* | 98.5 | 93.4 | 99.9 | 99.0 | 97.8 | 99.7 | 94.4 | 88.5 | 98.2 | 99.7 | 98.8 | 100.0 |
| *Proteus* | 96.9 | 84.9 | 99.9 | 99.3 | 98.3 | 99.9 | 88.9 | 74.1 | 97.8 | 99.8 | 99.1 | 100.0 |
| *Staphylococcus* | 99.3 | 96.4 | 100.0 | 95.6 | 93.2 | 97.5 | 84.5 | 76.4 | 91.0 | 99.8 | 99.1 | 100.0 |
| *Serratia* | 96.0 | 80.7 | 99.8 | 99.7 | 98.9 | 100.0 | 91.1 | 72.2 | 99.1 | 99.9 | 99.3 | 100.0 |

| **16S Metagenomics** | | | | | | | | | | | | |
|---|---|---|---|---|---|---|---|---|---|---|---|---|
| | **Sensitivity** | | | **Specificity** | | | **PPV** | | | **NPV** | | |
| **Target Genus** | **%** | **95% CI** | | **%** | **95% CI** | | **%** | **95% CI** | | **%** | **95% CI** | |
| *Acinetobacter* | 83.2 | 55.1 | 97.6 | 97.8 | 96.2 | 98.8 | 38.9 | 19.4 | 62.3 | 99.7 | 99.0 | 100.0 |
| *Escherichia* | 73.3 | 64.0 | 81.8 | 99.2 | 97.9 | 99.8 | 94.9 | 88.2 | 98.5 | 94.5 | 92.1 | 96.4 |
| *Enterobacter* | 21.1 | 15.3 | 34.9 | 86.5 | 83.2 | 89.4 | 10.6 | 6.1 | 18.6 | 93.7 | 90.8 | 95.9 |

| | | | | | | | | | | | | |
|---|---|---|---|---|---|---|---|---|---|---|---|---|
| *Haemophilus* | 85.2 | 74.8 | 93.0 | 88.0 | 84.6 | 90.8 | 50.5 | 40.5 | 60.6 | 97.6 | 95.6 | 99.0 |
| *Klebsiella* | 66.8 | 56.0 | 77.2 | 95.7 | 93.5 | 97.4 | 74.0 | 63.5 | 83.6 | 94.0 | 91.4 | 96.1 |
| *Moraxella* | 58.0 | 34.5 | 78.4 | 99.9 | 99.2 | 100.0 | 93.1 | 69.7 | 99.7 | 98.6 | 97.3 | 99.4 |
| *Pseudomonas* | 89.0 | 81.0 | 94.5 | 94.7 | 92.3 | 96.5 | 74.5 | 65.1 | 82.5 | 98.0 | 96.4 | 99.0 |
| *Proteus* | 54.6 | 36.1 | 71.7 | 99.9 | 99.2 | 100.0 | 95.5 | 77.4 | 99.8 | 97.6 | 96.0 | 98.7 |
| *Staphylococcus* | 82.8 | 74.3 | 89.2 | 77.9 | 73.8 | 81.8 | 47.4 | 40.2 | 55.0 | 95.0 | 92.1 | 96.9 |
| *Serratia* | 90.6 | 72.7 | 98.6 | 87.8 | 84.8 | 90.5 | 19.8 | 12.1 | 29.8 | 99.7 | 98.8 | 100.0 |

**Table S6.** Frequency of resistance gene detections by PCR tests among eligible samples (n = 606 for Unyvero, n = 620 for FilmArray)

| Resistance Gene Target | Unyvero | FilmArray |
|---|---|---|
| **Carbapenemases** | | |
| $bla_{IMP}$ | 0 | 1 |
| $bla_{KPC}$ | 1 | 1 |
| $bla_{OXA-23}$ | 5 | NA |
| $bla_{OXA24/40}$ | 0 | NA |
| $bla_{OXA-48}$ | 0 | 0 |
| $bla_{OXA-58}$ | 0 | NA |
| $bla_{NDM}$ | 2 | 0 |
| $bla_{VIM}$ | 3 | 1 |
| **Other beta-lactams** | | |
| $bla_{CTX-M}$ | 14 | 32 |

| | | |
|---|---|---|
| *bla*$_{SHV}$ | 55 | NA |
| *bla*$_{TEM}$ | 108 | NA |
| *mecA* | 92 | NA |
| *mecC* | 3 | NA |
| *mecA/C* and MREJ | NA | 32 |
| **Miscellaneous** | | |
| *ermB* | 68 | NA |
| *E. coli gyrA83* | 29 | NA |
| *P. aeruginosa gyrA87* | 35 | NA |
| *sul1* | 67 | NA |
